# Long-term Impact of Prior Secondhand Tobacco Smoke Exposure on Respiratory Health

**DOI:** 10.1101/2021.09.17.21263750

**Authors:** Fernando Diaz del Valle, Jonathan K. Zakrajsek, Sung-Joon Min, Patricia B. Koff, Harold W. Bell, Keegan A. Kincaid, Daniel N. Frank, Vijay Ramakrishnan, Moumita Ghosh, R. William Vandivier

## Abstract

**Rationale:** Ongoing secondhand tobacco smoke (SHS) exposure is associated with worsened respiratory health, but little is known about the long-term impact decades after exposure ended.

**Objective:** Determine the long-term consequences of SHS exposure on respiratory health.

**Methods:** Population-based, cohort study in subjects ≥50 years old who had >1 year *versus* ≤1 year of airline occupational SHS-exposure.

**Measurements and Main Results:** Respiratory health was the primary outcome measured by the St. George’s Respiratory Questionnaire (SGRQ). Key secondary outcomes included respiratory symptoms measured by COPD Assessment Test (CAT) and pre-bronchodilator lung function. The study enrolled 183 SHS-exposed and 59 unexposed subjects. SHS-exposed subjects were exposed to airline SHS for 16.1±9.3 years, which ended 27.5±9.4 years prior to enrollment. Prior SHS-exposure was associated with worse respiratory health based on a 6.7-unit increase in SGRQ (95% CI=[2.7, 10.7]; p=0.001) and 3-unit increase in CAT (95% CI=1.4, 4.6]; p<0.001) *versus* unexposed subjects, but was not associated with airflow obstruction defined by FEV_1_:FVC<0.7. Clinically-significant respiratory symptoms (CAT≥10) in SHS-exposed never smokers with preserved lung function (FEV_1_:FVC ≥0.7 and FVC ≥lower limit of normal) were associated with decreased respiratory and non-respiratory quality-of-life, reduced lung function that remained within the normal range, increased comorbidities and inhaled bronchodilator use, higher plasma CRP and SAA-1 and fewer sinonasal basal stem/progenitor cells *versus* asymptomatic (CAT<10) SHS-exposed subjects.

**Conclusion:** SHS is associated with a phenotype of impaired respiratory health almost 3 decades after exposure ended, consistent with a symptomatic form of COPD with preserved lung function recently described in smokers.

## INTRODUCTION

The World Health Organization estimates that 1.8 billion people worldwide continue to be exposed to SHS, resulting in more than 600,000 deaths per year(1, 2). In the United States, SHS-exposure affects 58 million people annually, many of whom are children, communities of color, or people living in poverty or rental housing(3, 4). SHS exposure in the U.S. also causes over 42,000 deaths every year, including 900 infants(4). This common exposure has well-established, serious consequences for cardiovascular disease, stroke, lung cancer, sudden infant death syndrome, reproductive health and respiratory health, especially for children(5, 6). In adults, ongoing SHS exposure is associated with increased respiratory symptoms and a small decrement in lung function, but its long-term impact on respiratory health and the development of obstructive and non-obstructive forms of chronic obstructive pulmonary disease (COPD) decades after exposure has stopped are uncertain(5, 6).

COPD is a major cause of chronic respiratory symptoms and poor quality of life for millions of people(7). Prolonged exposure to inhaled particulates, such as tobacco smoke, cause chronic inflammation that narrows or obliterates airways, destroys lung tissue and increases airway mucus production in susceptible people(8-10). These pathologic changes impair lung growth, shorten the plateau phase of lung function and accelerate lung function decline(11-13), leading to fixed expiratory airflow obstruction that is the *sine qua non* of classical COPD(7). Recent studies also show that smoking can cause a symptomatic form of COPD with preserved lung function in up to 50% of smokers and former smokers(14-21). These smokers have a phenotype of respiratory symptoms, chronic bronchitis, increased respiratory medication use, exacerbations, thickened airways, small decrements in lung function within the normal range (*e*.*g*. FEV_1_:FVC ratio ≥ 0.7) and systemic inflammation(14-22), suggesting that they have a non-classical form of symptomatic COPD with preserved lung function.

Globally, ∼25-45% of COPD is not directly caused by smoking, but instead is thought to result from environmental exposures, such as SHS(23-29). Despite this potential impact, the long-term effects of SHS on respiratory health remain unresolved, especially as it relates to non-classical forms of COPD with preserved lung function. Accordingly, this study was designed to test the hypothesis that SHS exposure is associated with poor respiratory health decades after exposure has ceased, and that the presence of respiratory symptoms in SHS-exposed subjects with preserved lung function has clinical and biologic consequences (Fig. 1A). This hypothesis was tested in flight attendants because of their long, well-documented exposure to SHS that stopped over 20 years ago, giving the opportunity to examine the long-term effects of SHS on respiratory health without ongoing exposure (Fig. 1B)(30).

**Figure 1.**
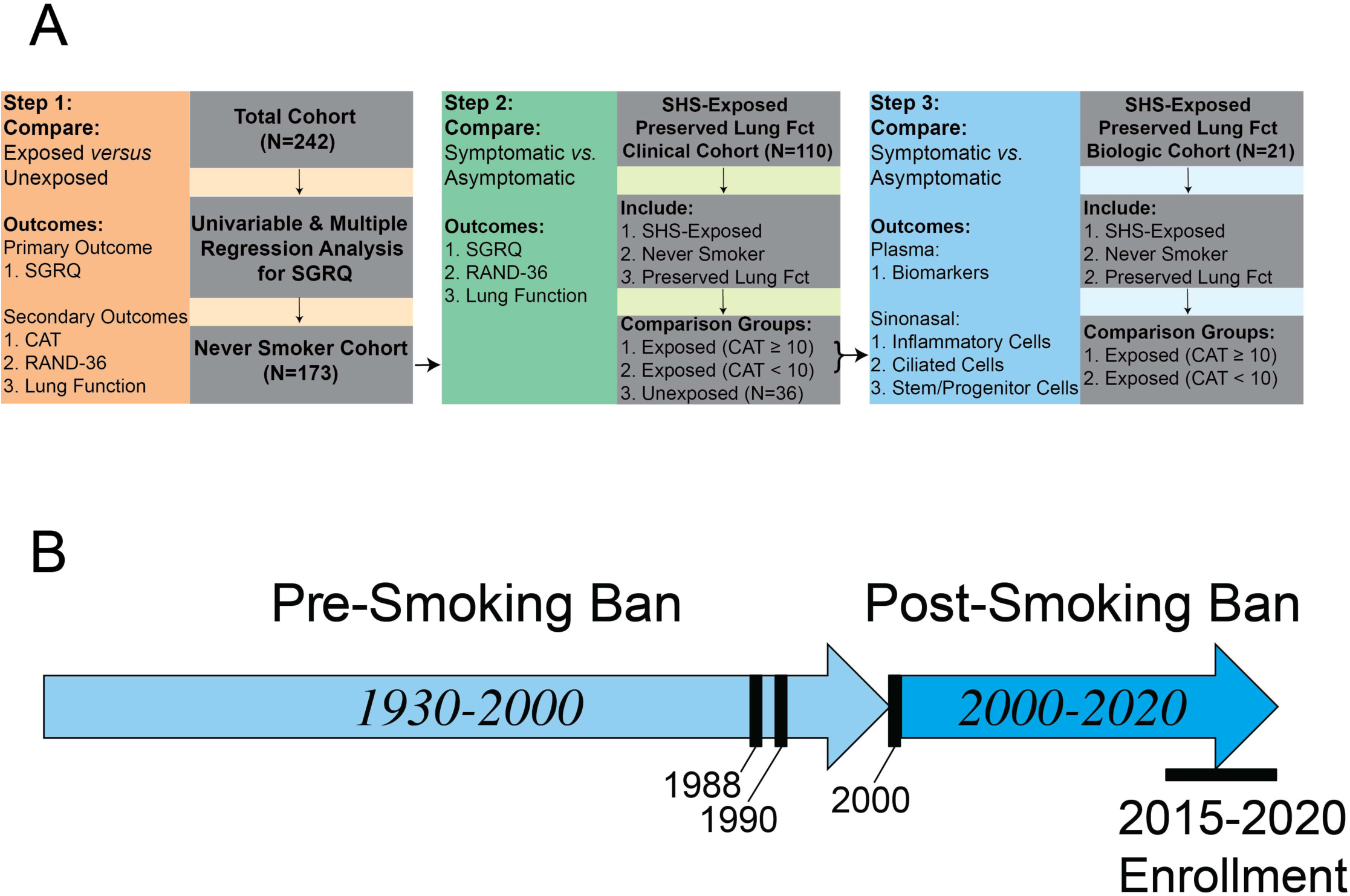
Study Design and Flight Attendant Secondhand Tobacco Smoke (SHS) Exposure History. A) Study Design for main (Step 1) and secondary analyses (Steps 2 and 3). Abbreviations: SGRQ – St. George’s Respiratory Questionnaire; CAT – COPD Assessment Test; RAND-36 – RAND Corporation modification of the short form (SF)-36 questionnaire; Fct – function. B) Flight attendants were involuntarily exposed to SHS while working for the airlines from 1930 to 2000 (light blue arrow). National smoking bans were enacted in 1988 (2 hour domestic flights), 1990 (6 hour domestic flights) and 2000 (all domestic and international flights), which are indicated by the vertical black bars. Flight attendants have not been exposed to airline SHS since 2000, indicated by the dark blue arrow. Flight attendants were enrolled and measurements were taken during the period from 2015-2020, indicated by the horizontal black bar.

## METHODS

### Study Design

We conducted a cohort study from 2015 through 2020. The Colorado Multiple Institutional Review Board approved the study (COMIRB 14-0556) and all subjects signed informed consent. De-identified data from this study will be made available to other investigators upon request.

### Study Population

Subjects were recruited from flight attendant organizations and Facebook groups. Participants recruited were all ≥ 50 years of age. The SHS-exposed group (*i*.*e*. exposed) worked as flight attendants for ≥ 1 year where they were exposed to SHS for > 1 year. The SHS-unexposed group (*i*.*e*. unexposed) worked as a flight attendant for ≥ 1 year where they were exposed to SHS for ≤ 1 year, or never worked as a flight attendant. The 1-year cutoff for SHS exposure was based on data demonstrating that exposure to particulates for at least one year had a stronger effect on lung function than shorter-term exposure (reference 5, page 554)(5, 31). Subjects were excluded for smoking within the previous year, pregnancy, treatment for cancer, prior surgical lung procedures or inability to understand the consent.

### Measurements

#### Clinical Measurements

All measurements were taken at the time of enrollment from 2015-2020, which was ≥15 years after their last possible exposure to airline SHS (Fig. 1B)(30). Information about demographics, comorbidities and current medications was collected from all subjects at enrollment. Participants answered the tobacco smoke exposure questionnaire modeled after Eisner *et al*.*(32)* and adapted from Arjomandi *et al*.(33), which collected detailed smoking and SHS-exposure history, including pre-natal, home, adult non-airline work and adult airline work exposure. The questionnaire also collected information about exposures to vapors, gas, dusts, and fumes (VGDF). Subjects reported current use of inhaled corticosteroids (ICS), and inhaled bronchodilators, including short-acting β2 agonists (SABA), long-acting β2 agonists (LABA), and long-acting anti-muscarinics (LAMA). *Primary and Secondary Outcomes*. The primary study outcome was respiratory quality-of-life, measured by the total SGRQ score(34). Key secondary outcomes included the CAT(35) which measured respiratory symptoms, the Rand Corporation modification of the 36-item Short Form (SF) Health Survey Questionnaire (RAND-36)(36) which measure general quality of life, and pre-bronchodilator (BD) spirometry including FEV_1_, FVC and FEV_1_:FVC ratio (ndd Medical Technologies, Andover, MA 01810). Post-BD spirometry, lung diffusion of carbon monoxide (DLCO), and lung volumes by the nitrogen washout method were performed on a subset of subjects. The study used reference equations by Quanjer *et al*. for spirometry(37) and Crapo *et al*. for lung volumes and DLCO(38).

#### Measurement of Plasma Biomarkers

The Human Biomarker 46-Vplex (Meso Scale Diagnostics, Rockville, MD 20850) and NT-ProBNP kit (Abcam, Cambridge, MA 02139) were used to measure plasma mediators in a subset of subjects.

#### Nasal Brushings

Nasal brushings were performed in a subset of subjects using published methods(39). Cells were plated by Cytospin, stained with Diff-Quick, or fixed in 10% neutral-buffered formalin and stored in 0.1 % sodium azide at 4°C for later immunostaining.

#### Immunofluorescence

Immunostaining of cytospins was performed with basal stem/progenitor markers keratin (K) 5 and p63 following standard protocols as previously published(40). Images were acquired using an upright Zeiss Imager Z1 Fluorescent microscope and Zen 2 (blue edition) software (Carl Zeiss AG, White Plains, NY).

### Statistics

Differences between SHS-exposed and unexposed groups in demographics, comorbidities, primary or secondary outcomes were determined using t-tests (or Mann-Whitney tests if skewed) for continuous variables and chi-square tests (or Fisher’s exact tests if subject numbers were small) for categorical variables. Differences in these same variables were also assessed in the never-smoker cohort, defined as a pack-year smoking history of ≤1 year(14, 17). Differences by group were estimated using quantile regression (for medians) for selected skewed variables. Additional post-hoc analyses included: 1) Effect of SHS-exposure on total SGRQ adjusting for variables in regression, 2) Synergistic interaction between SHS-exposure and former smoking on total SGRQ, total CAT, and obstruction using regression to assess the interaction effect.

The impact of symptoms on clinical and biologic outcomes was examined in three groups of never-smokers with preserved lung function, defined as FEV_1_:FVC ratio ≥ 0.7 and FVC ≥ lower limit of normal using post-BD measurements where available(14, 37): 1) Symptomatic (*i*.*e*. CAT ≥ 10) SHS-exposed, 2) Asymptomatic (*i*.*e*. CAT < 10) SHS-exposed, and 3) SHS-unexposed. Differences between these groups were assessed using analysis of variance (ANOVA) with Dunnett’s tests or Kruskal-Wallis tests with Dunn’s tests. Differences in SGRQ were also examined after removing subjects with a prior history of asthma. T-tests were used to examine differences in plasma mediators and sinonasal airway basal stem/progenitor cells between symptomatic *versus* asymptomatic, SHS-exposed, never-smokers with preserved lung function. Pearson correlation coefficients were estimated to assess the association between airway stem/progenitors and total CAT, CRP and total lung capacity (TLC). All tests were 2-sided with a significance level of p<0.05. SAS v.9.4 (SAS Institute, Cary, NC) and GraphPad Prism Version 9.0.0 (San Diego, CA) were used for all analyses.

### Power Analysis

The power analysis assumed that the exposed group would have a mean total SGRQ of 15.2 with a standard deviation (SD) of 15.6. Based on 2:1 enrollment of exposed and unexposed subjects, respectively, and a 5% dropout rate, the study would have 80% power to detect a 4-unit minimum clinically important difference in the total SGRQ by enrolling 380 subjects in the exposed group and 190 in the unexposed group(41). With the same criteria 80% power to detect a 5-unit difference in total SGRQ will need enrolling 242 exposed and 121 unexposed, and a 6-unit difference from 168 exposed and 84 unexposed subjects.

## RESULTS

The study enrolled 242 subjects, including 183 exposed and 59 unexposed. Exposed subjects were older, more likely to be female, and had a lower BMI, compared to unexposed subjects (Table 1) Exposed subjects also reported less non-airline work SHS-exposure and more exposure to VGDF in the workplace (Table 1), compared to unexposed subjects (Table 1). Exposed subjects were hired as a flight attendant at a mean age of 23.8 ± 4.8 years, exposed to aircraft SHS for 16.1 ± 9.3 years and stopped being exposed 27.5 ± 9.4 years prior to study enrollment. There were no differences in race/ethnicity, past smoking history or comorbidities (Table 1).

**Table 1.**
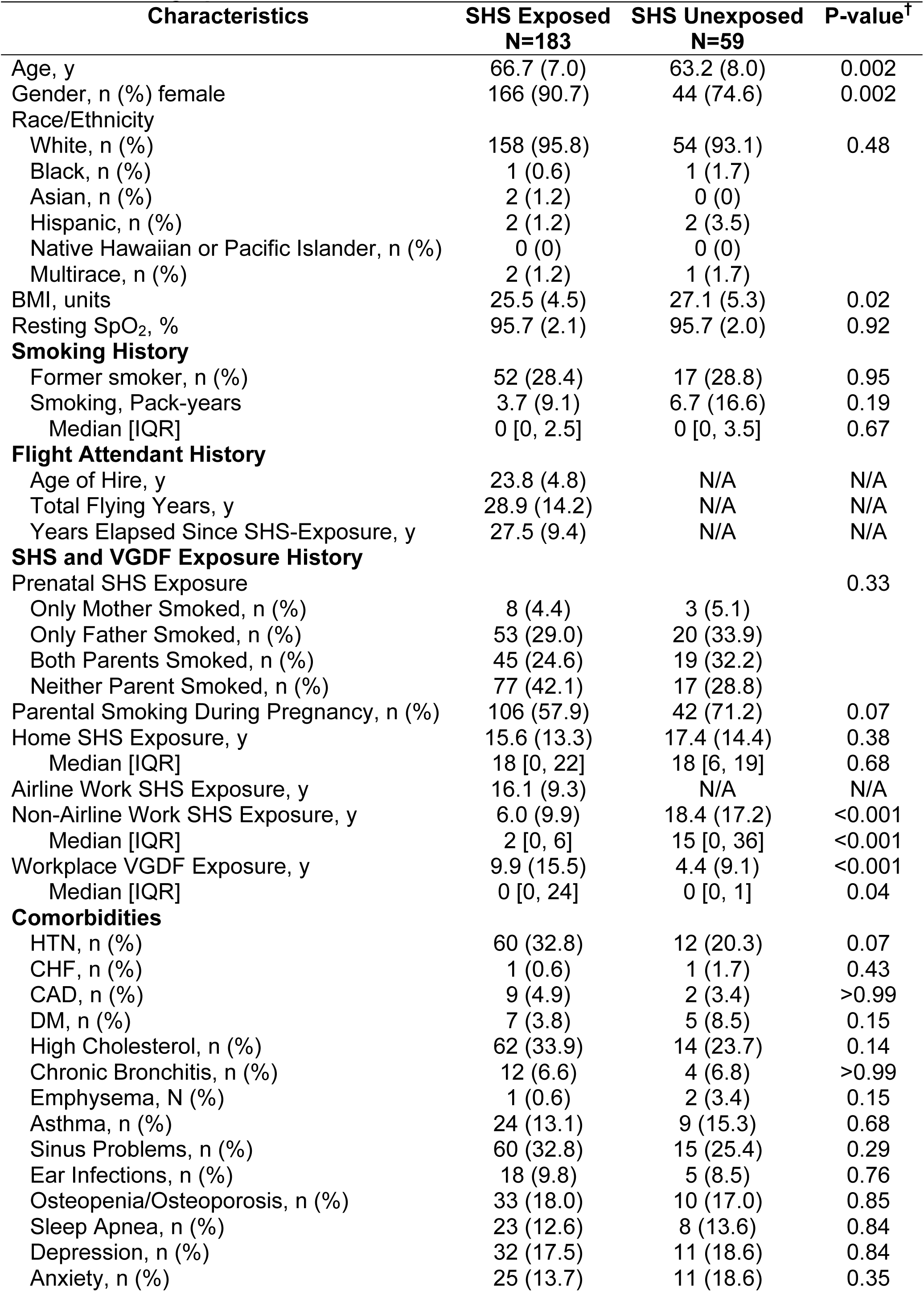
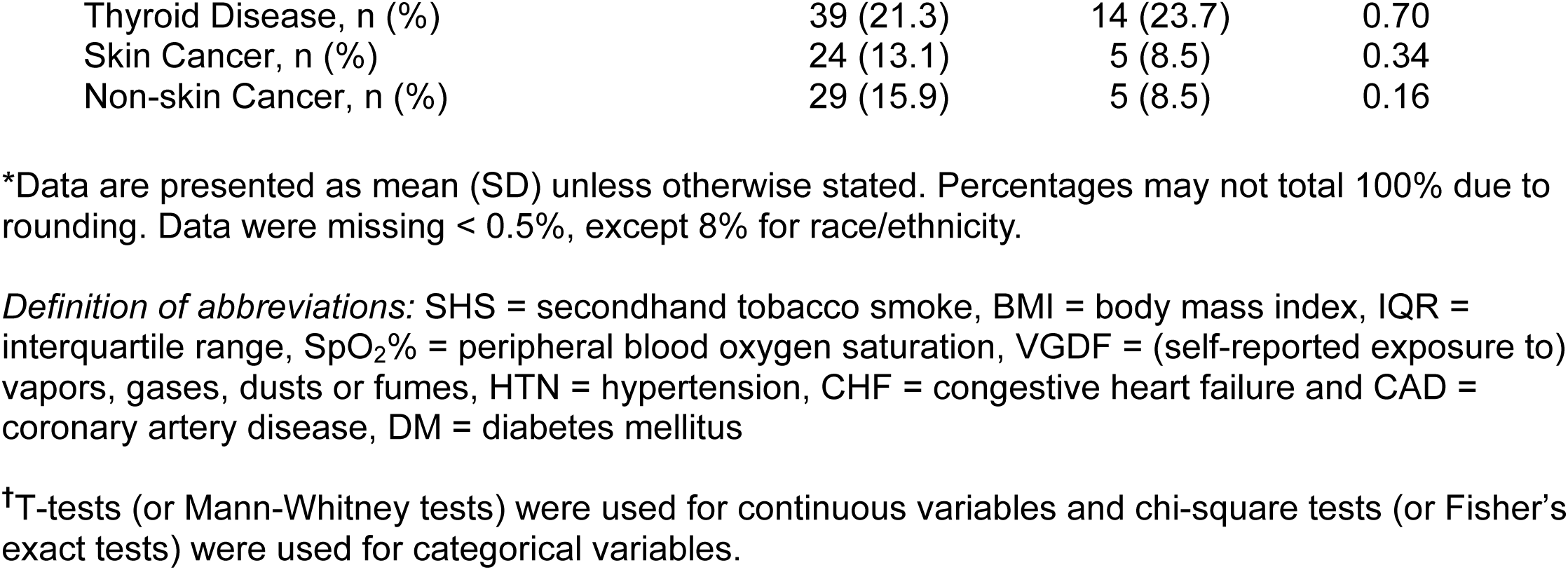
Demographics of Total Cohort*.

| Characteristics | SHS Exposed<br>N=183 | SHS Unexposed<br>N=59 | P-value<sup>†</sup> |
| --- | --- | --- | --- |
| Age, y | 66.7 (7.0) | 63.2 (8.0) | 0.002 |
| Gender, n (%) female | 166 (90.7) | 44 (74.6) | 0.002 |
| Race/Ethnicity |  |  |  |
| White, n (%) | 158 (95.8) | 54 (93.1) | 0.48 |
| Black, n (%) | 1 (0.6) | 1 (1.7) |  |
| Asian, n (%) | 2 (1.2) | 0 (0) |  |
| Hispanic, n (%) | 2 (1.2) | 2 (3.5) |  |
| Native Hawaiian or Pacific Islander, n (%) | 0 (0) | 0 (0) |  |
| Multirace, n (%) | 2 (1.2) | 1 (1.7) |  |
| BMI, units | 25.5 (4.5) | 27.1 (5.3) | 0.02 |
| Resting SpO<sub>2</sub>, % | 95.7 (2.1) | 95.7 (2.0) | 0.92 |
| **Smoking History** |  |  |  |
| Former smoker, n (%) | 52 (28.4) | 17 (28.8) | 0.95 |
| Smoking, Pack-years | 3.7 (9.1) | 6.7 (16.6) | 0.19 |
| Median [IQR] | 0 [0, 2.5] | 0 [0, 3.5] | 0.67 |
| **Flight Attendant History** |  |  |  |
| Age of Hire, y | 23.8 (4.8) | N/A | N/A |
| Total Flying Years, y | 28.9 (14.2) | N/A | N/A |
| Years Elapsed Since SHS-Exposure, y | 27.5 (9.4) | N/A | N/A |
| **SHS and VGDF Exposure History** |  |  |  |
| Prenatal SHS Exposure |  |  | 0.33 |
| Only Mother Smoked, n (%) | 8 (4.4) | 3 (5.1) |  |
| Only Father Smoked, n (%) | 53 (29.0) | 20 (33.9) |  |
| Both Parents Smoked, n (%) | 45 (24.6) | 19 (32.2) |  |
| Neither Parent Smoked, n (%) | 77 (42.1) | 17 (28.8) |  |
| Parental Smoking During Pregnancy, n (%) | 106 (57.9) | 42 (71.2) | 0.07 |
| Home SHS Exposure, y | 15.6 (13.3) | 17.4 (14.4) | 0.38 |
| Median [IQR] | 18 [0, 22] | 18 [6, 19] | 0.68 |
| Airline Work SHS Exposure, y | 16.1 (9.3) | N/A | N/A |
| Non-Airline Work SHS Exposure, y | 6.0 (9.9) | 18.4 (17.2) | <0.001 |
| Median [IQR] | 2 [0, 6] | 15 [0, 36] | <0.001 |
| Workplace VGDF Exposure, y | 9.9 (15.5) | 4.4 (9.1) | <0.001 |
| Median [IQR] | 0 [0, 24] | 0 [0, 1] | 0.04 |
| **Comorbidities** |  |  |  |
| HTN, n (%) | 60 (32.8) | 12 (20.3) | 0.07 |
| CHF, n (%) | 1 (0.6) | 1 (1.7) | 0.43 |
| CAD, n (%) | 9 (4.9) | 2 (3.4) | >0.99 |
| DM, n (%) | 7 (3.8) | 5 (8.5) | 0.15 |
| High Cholesterol, n (%) | 62 (33.9) | 14 (23.7) | 0.14 |
| Chronic Bronchitis, n (%) | 12 (6.6) | 4 (6.8) | >0.99 |
| Emphysema, N (%) | 1 (0.6) | 2 (3.4) | 0.15 |
| Asthma, n (%) | 24 (13.1) | 9 (15.3) | 0.68 |
| Sinus Problems, n (%) | 60 (32.8) | 15 (25.4) | 0.29 |
| Ear Infections, n (%) | 18 (9.8) | 5 (8.5) | 0.76 |
| Osteopenia/Osteoporosis, n (%) | 33 (18.0) | 10 (17.0) | 0.85 |
| Sleep Apnea, n (%) | 23 (12.6) | 8 (13.6) | 0.84 |
| Depression, n (%) | 32 (17.5) | 11 (18.6) | 0.84 |
| Anxiety, n (%) | 25 (13.7) | 11 (18.6) | 0.35 |
| Thyroid Disease, n (%) | 39 (21.3) | 14 (23.7) | 0.70 |
| Skin Cancer, n (%) | 24 (13.1) | 5 (8.5) | 0.34 |
| Non-skin Cancer, n (%) | 29 (15.9) | 5 (8.5) | 0.16 |

\*Data are presented as mean (SD) unless otherwise stated. Percentages may not total 100% due to rounding. Data were missing < 0.5%, except 8% for race/ethnicity.
*Definition of abbreviations:* SHS = secondhand tobacco smoke, BMI = body mass index, IQR = interquartile range, SpO<sub>2</sub>% = peripheral blood oxygen saturation, VGDF = (self-reported exposure to) vapors, gases, dusts or fumes, HTN = hypertension, CHF = congestive heart failure and CAD = coronary artery disease, DM = diabetes mellitus
†T-tests (or Mann-Whitney tests) were used for continuous variables and chi-square tests (or Fisher's exact tests) were used for categorical variables.

SHS-exposure was associated with a significant reduction in respiratory quality of life, measured by a 6.7 unit clinically significant increase in the median total SGRQ score (95% Confidence Interval [CI] = [2.7, 10.7]; p=0.001) in the exposed *versus* unexposed groups (Fig. 2A)(41). Symptom, Activity and Impact component scores of the SGRQ were also higher (*i*.*e*. worse) in the exposed *versus* unexposed groups (Fig. 2B). Respiratory symptoms were higher in exposed *versus* unexposed subjects, as assessed by the total CAT (3 unit difference of medians; 95% CI = [1.4, 4.6]; p<0.001) and CAT respiratory component score (Fig. 2C and E). The difference in total CAT was above the 2 unit minimum clinically important difference(42). SHS-exposed subjects were more likely to have clinically significant respiratory symptoms, defined as a total CAT ≥ 10 or a CAT respiratory component score ≥ 7 (Fig. 2D and F)(7, 43). General quality of life, measured by the RAND-36 physical function and social function, were lower (*i*.*e*. worse) in the exposed *versus* unexposed groups (Supplement Fig. 1A-B). In contrast, exposure had a varying effect on pre- and post-BD spirometry (Fig. 2G-I and Supplement Fig. 1C-E), lung volumes and DLCO (Supplement Fig. 1F-G), and was not associated with more obstruction defined as an FEV_1_:FVC ratio < 0.7 (Fig. 2J)(7). The exposed group also did not use more SABA, LABA, LAMA, or ICS inhaler medications (data not shown).

**Figure 2.**
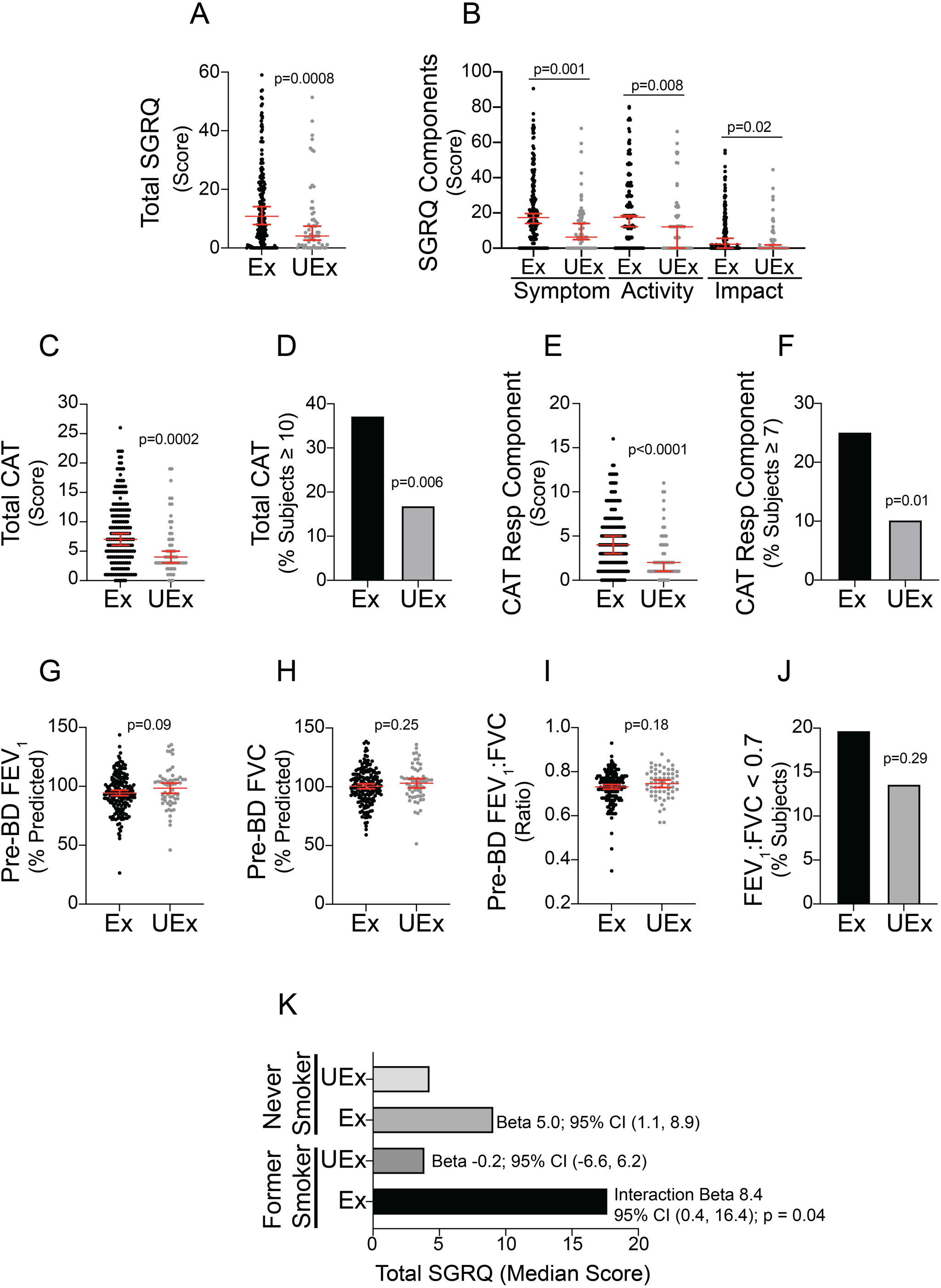
Impact of Secondhand Tobacco Smoke (SHS)-Exposure on Respiratory Health in the Total Cohort. The study examined differences in SHS-exposed (Ex; N=183) *versus* unexposed (UEx; N=59) subjects for A) total St. George’s Respiratory Questionnaire (SGRQ), B) symptom, activity and impact components of the SGRQ, C) total COPD Assessment Test (CAT), D) total CAT ≥ 10, E) CAT Respiratory (Resp) Component, F) CAT Resp Component ≥ 7, pre-bronchodilator (pre-BD) G) FEV_1_, H) FVC, I) FEV_1_:FVC ratio, and J) obstruction defined by an FEV_1_:FVC ratio < 0.7 using post-BD measurements where available. K) The study examined whether SHS-exposure and former smoking interacted in a synergistic way for total SGRQ compared to UEx never smokers. P-values indicate results of a Mann-Whitney test for A-C and E, and data are shown as median and 95% CI. P-values indicate results of a t-test for G-I, and data are shown as mean and 95% CI. P-values indicate results of a Fisher’s exact test for D, F and J. Beta and 95% CI indicate the main and interaction effects of Ex and former smoking using quantile regression for K.

Univariable analysis demonstrated that total SGRQ was not affected by female gender, parental smoking during pregnancy, exposure to VGDF in the workplace or hypertension, but was affected by age, BMI, airline work SHS-exposure and marginally by non-airline work SHS-exposure (Supplement Table 1). The multivariable model showed that total SGRQ was higher in the exposed *versus* unexposed groups (beta=6.5; 95% CI = [3.7, 9.2]; p<0.001), when adjusted for BMI and parental smoking during pregnancy, which were selected using a backward elimination algorithm (Supplement Table 1). Interestingly, SHS-exposure interacted synergistically with former smoking to increase total SGRQ (Fig. 2K), but not total CAT or obstruction defined as an FEV_1_:FVC ratio < 0.7 (Supplement Fig. 4A-B). A sensitivity analysis performed in never-smokers mirrored results in the total cohort (Supplement Table 2, Supplement Fig. 2 and 3).

### Impact of Symptoms in the SHS-Exposed, Preserved Lung Function Cohort

We sought to determine whether the presence of symptoms in SHS-exposed subjects with preserved lung function had important clinical and biological consequences. The analysis was restricted to never-smokers to eliminate any potential confounding effect of past smoking.

Respiratory symptoms, defined as a total CAT ≥ 10, were present in 33% of SHS-exposed subjects with preserved lung function (Table 2). Demographic characteristics of symptomatic (*i*.*e*. CAT ≥ 10) and asymptomatic (*i*.*e*. CAT < 10) subjects differed in that the presence of symptoms was associated with a higher BMI, more parental smoking during pregnancy, more home SHS exposure, and an increased prevalence of chronic bronchitis, depression, anxiety and a trend for asthma (Table 2).

**Table 2.** Demographics of Never Smokers with Preserved Lung Function^*,†^.

| Characteristics | SHS Exposed<br>CAT≥10<br>N=36 | SHS Exposed<br>CAT<10<br>N=74 | P-value‡ |
| --- | --- | --- | --- |
| Age, y | 65.6 (8.6) | 65.1 (6.7) | 0.77 |
| Gender, n (%) female | 34 (94.4) | 68 (91.9) | >0.99 |
| Race/Ethnicity |  |  |  |
| White, n (%) | 32 (97.0) | 64 (94.1) | >0.99 |
| Black, n (%) | 0 (0) | 0 (0) |  |
| Asian, n (%) | 0 (0) | 2 (2.9) |  |
| Hispanic, n (%) | 0 (0) | 2 (2.9) |  |
| Native Hawaiian or Pacific Islander, n (%) | 0 (0) | 0 (0) |  |
| Multirace, n (%) | 1 (3.0) | 0 (0) |  |
| BMI, units | 26.8 (5.0) | 24.6 (3.6) | 0.02 |
| Resting SpO<sub>2</sub>, % | 95.3 (2.2) | 95.9 (2.0) | 0.19 |
| **Flight Attendant History** |  |  |  |
| Age of Hire, y | 23.0 (3.8) | 24.2 (4.6) | 0.17 |
| Total Flying Years, y | 30.5 (13.6) | 28.8 (14.6) | 0.57 |
| Total Flying SHS Exposure, y | 17.2 (10.9) | 14.2 (8.2) | 0.14 |
| Years Elapsed Since SHS-Exposure, y | 26.2 (9.1) | 27.7 (8.8) | 0.44 |
| **SHS and VGDF Exposure History** |  |  |  |
| Prenatal SHS Exposure |  |  |  |
| Only Mother Smoked, n (%) | 2 (5.6) | 4 (5.4) | 0.13 |
| Only Father Smoked, n (%) | 14 (38.9) | 17 (23.0) |  |
| Both Parents Smoked, n (%) | 9 (25.0) | 13 (17.6) |  |
| Neither Parent Smoked, n (%) | 11 (30.6) | 40 (54.1) |  |
| Parental Smoking During Pregnancy, n (%) | 25 (69.4) | 34 (46.0) | 0.02 |
| Home SHS Exposure, y | 15.3 (12.2) | 10.9 (10.3) | 0.047 |
| Median [IQR] | 19 [0, 22.5] | 13.5 [0, 18] | 0.02 |
| Airline Work SHS Exposure, y | 17.2 (10.9) | 14.2 (8.2) | 0.14 |
| Non-Airline Work SHS Exposure, y | 5.6 (9.8) | 4.0 (7.2) | 0.39 |
| Median [IQR] | 0 [0, 7.5] | 1.3 [0, 4] | 0.86 |
| Workplace VGDF Exposure, y | 12.2 (15.6) | 10.8 (16.1) | 0.67 |
| Median [IQR] | 0 [0, 28] | 0 [0, 27] | 0.59 |
| **Comorbidities** |  |  |  |
| HTN, n (%) | 7 (19.4) | 24 (32.4) | 0.16 |
| CHF, n (%) | 0 (0) | 0 (0) | N/A |
| CAD, n (%) | 1 (2.8) | 3 (4.1) | >0.99 |
| DM, n (%) | 0 (0) | 2 (2.7) | >0.99 |
| High Cholesterol, n (%) | 14 (38.9) | 18 (24.3) | 0.11 |
| Chronic Bronchitis, n (%) | 5 (13.9) | 0 (0) | 0.003 |
| Emphysema, n (%) | 0 (0) | 0 (0) | N/A |
| Asthma, n (%) | 6 (16.7) | 3 (4.1) | 0.06 |
| Sinus Problems, n (%) | 14 (38.9) | 19 (25.7) | 0.16 |
| Ear Infections, n (%) | 4 (11.1) | 8 (10.8) | >0.99 |
| Osteopenia/Osteoporosis, n (%) | 6 (16.7) | 14 (18.9) | 0.77 |
| Sleep Apnea, n (%) | 5 (13.9) | 4 (5.4) | 0.15 |
| Depression, n (%) | 9 (25.0) | 6 (8.1) | 0.02 |
| Anxiety, n (%) | 10 (27.8) | 9 (12.2) | 0.04 |
| Thyroid Disease, n (%) | 7 (19.4) | 14 (18.9) | 0.95 |
| Skin Cancer, n (%) | 4 (11.1) | 9 (12.2) | >0.99 |
| Total Non-skin Cancer, n (%) | 6 (16.7) | 10 (13.5) | 0.66 |

\*Preserved lung function defined at an FEV<sub>1</sub>:FVC ratio $\geq 0.7$ and FVC ≥ lower limit of normal.
†Data are presented as mean (SD) unless otherwise stated. Percentages may not total 100% due to rounding. Data were missing < 1%, except 8% for race/ethnicity.
*Definition of abbreviations:* CAT = COPD Assessment Test, SHS = secondhand tobacco smoke, BMI = body mass index, IQR = interquartile range, SpO<sub>2</sub>% = peripheral blood oxygen saturation, VGDF = (self-reported exposure to) vapors, gases, dusts or fumes, HTN = hypertension, CHF = congestive heart failure and CAD = coronary artery disease, DM = diabetes mellitus
‡T-tests (or Mann-Whitney tests) were used for continuous variables and chi-square tests (or Fisher's exact tests) were used for categorical variables.

Symptomatic subjects had worse respiratory quality of life (*i*.*e*. total SGRQ and SGRQ components; Fig. 3A-D), general quality of life (*i*.*e*. all RAND-36 components; Fig. 3E-L) as well as a decrement in pre- and post-BD FEV_1_, FVC and TLC (Fig. 3M-N and Supplement Fig. 5A, B and D), compared to asymptomatic subjects or to unexposed controls (N=36). There were no differences in pre- or post-BD FEV_1_:FVC ratio (Fig. 3O and Supplement Fig. 5C) or DLCO (Supplement Fig. 5E). Symptomatic subjects were more likely to use inhaled bronchodilators compared to asymptomatic subjects (16.7 vs. 1.4%; p=0.005). The impact of symptoms on total SGRQ persisted when subjects with a past history of asthma were removed (14.0 unit difference of total SGRQ medians; 95% CI = [7.0, 21.0]; p<0.001). Therefore, the presence of respiratory symptoms in SHS-exposed subjects with preserved lung function identified a subgroup with clinically important consequences, compared to asymptomatic subjects.

**Figure 3.**
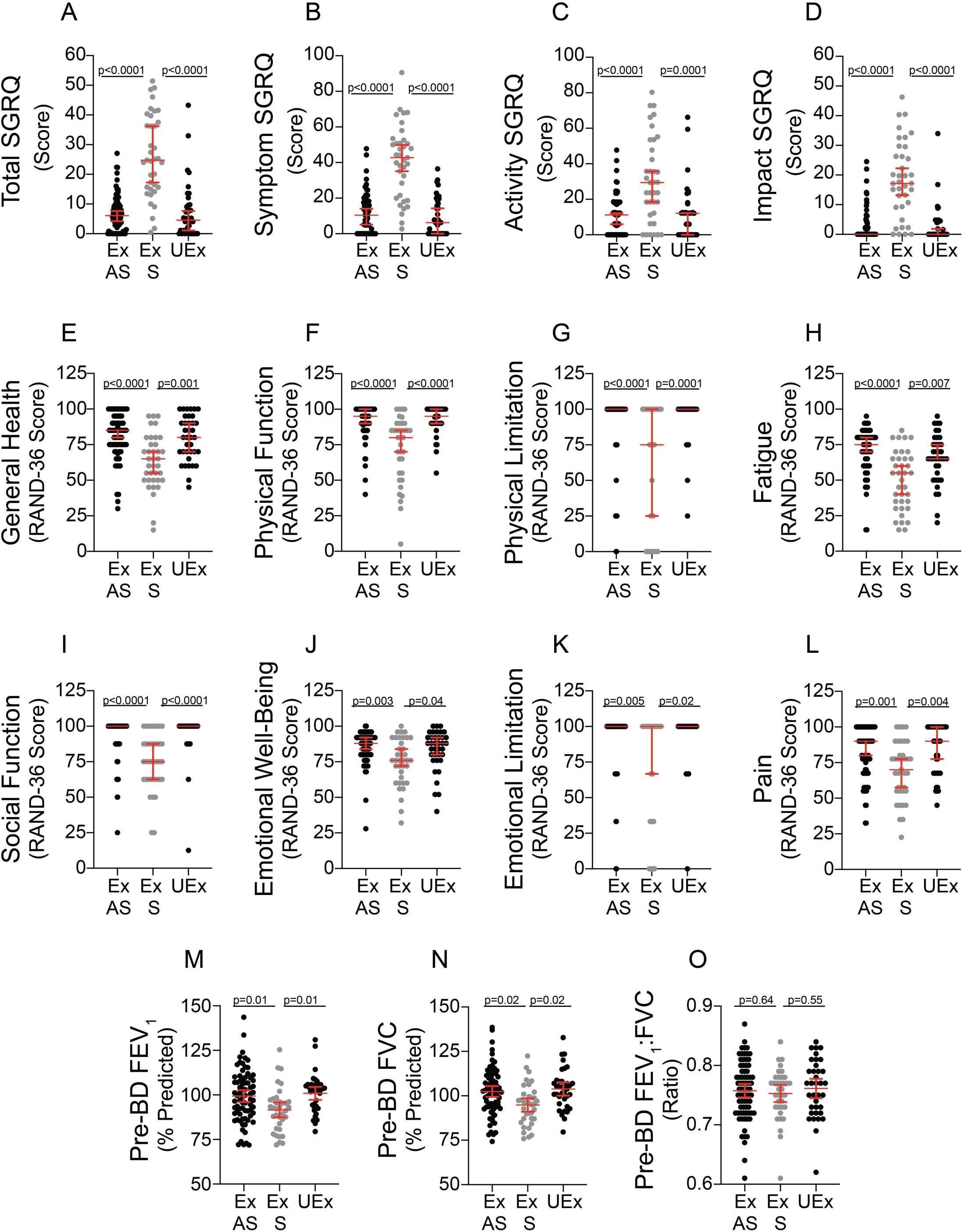
Clinical Impact of Symptoms in the Secondhand Tobacco Smoke (SHS) Exposed, Preserved Lung Function Cohort. The study examined the impact of symptoms on clinical outcomes in SHS-exposed (Ex), never-smokers with preserved lung function, defined as FEV_1_:FVC ratio ≥ 0.7 and FVC ≥ lower limit of normal. Symptomatic (S; CAT ≥ 10) SHS-exposed subjects with preserved lung function (N=36) were compared to asymptomatic (AS; CAT < 10) SHS-exposed subjects (N=74) and to SHS-unexposed (UEx) never-smokers with preserved lung function (N=36) for A) total, B) symptom, C) activity, and D) impact components of the St. George’s Respiratory Questionnaire (SGRQ), E) general health, F) physical function, G) role limitation due to physical health (physical limitation), H) fatigue, I) social function, J) emotional well-being, K) role limitation due to emotional problems (emotional limitation) and L) pain components of the RAND Corporation modification of the short form (SF)-36 questionnaire (RAND-36), and pre-bronchodilator (pre-BD) M) FEV_1_, N) FVC and O) FEV_1_:FVC ratio. Differences were assessed using a Kruskal-Wallis test for A-L, which resulted in a p < 0.006 for A-L. P-values indicate results of a Dunns test and data are shown as median and 95% CI. Differences were assessed using a one-way ANOVA for M-O, which resulted in a p < 0.02 for M-N. P-values indicate results of a Dunnett’s test and data are shown as mean and 95% CI.

We examined whether symptoms were associated with a biologic impact in SHS-exposed, never-smokers with preserved lung function by measuring plasma inflammatory mediators (Fig 4A-F), sinonasal airway basal stem/progenitor cells (Fig. 4G-H), as well as sinonasal inflammatory and ciliated cells (Supplement Fig. 6A-C). The mean plasma CRP and SAA-1 were increased in symptomatic *versus* asymptomatic subjects (Fig. 4A-B). IL-21 and ICAM-1 trended toward being significantly elevated in symptomatic subjects, whereas VCAM-1 and NT-ProBNP did not (Fig. 4C-F). The mean percentage of K5/p63 positive sinonasal airway basal stem/progenitor cells was reduced in sysmptomatic *versus* asymptomatic subjects (Fig. 4G-H). The percent of basal stem/progenitor cells correlated negatively with symptoms measured by the total CAT (Fig. 4I). They also correlated negatively with plasma CRP (Fig. 4J) and positively with TLC by nitrogen washout (Fig. 4K), but did not reach statistical significance. In contrast, symptoms did not affect the mean percentage of inflammatory or ciliated epithelial cells in nasal brushings (Supplement Fig. 6A-C),

**Figure 4.**
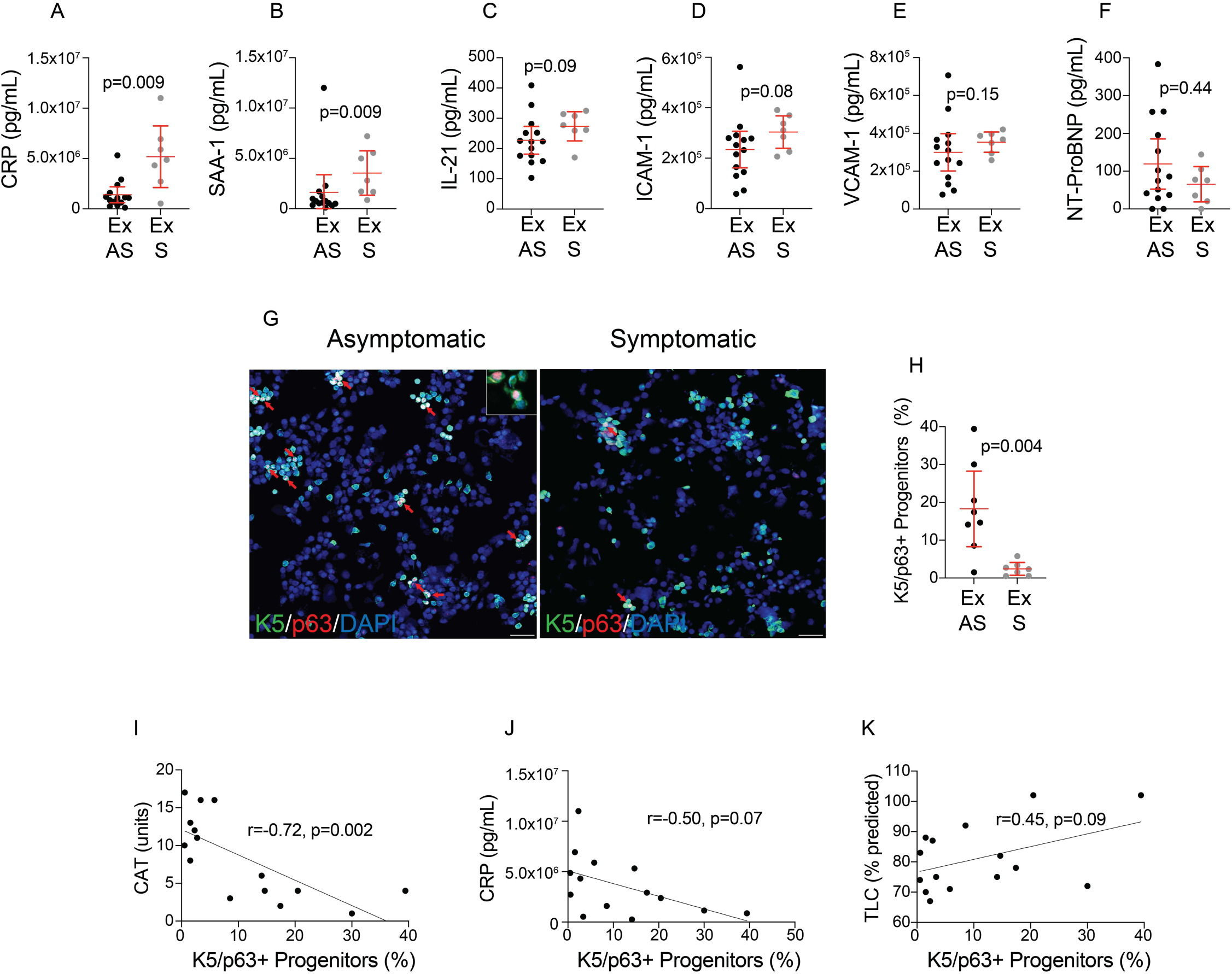
Biologic Impact of Symptoms in the Secondhand Tobacco Smoke (SHS) Exposed, Preserved Lung Function Cohort. The study examined the impact of symptoms on biologic outcomes in SHS-exposed (Ex), never-smokers with preserved lung function. Symptomatic (S; CAT ≥ 10; N=7) SHS-exposed subjects with preserved lung function were compared to asymptomatic (AS; CAT < 10; N=14) SHS-exposed subjects for plasma A) CRP, B) SAA-1, C) IL-21, D) ICAM-1 E) VCAM-1 and F) NT-ProBNP. Nasal brushing were also collected from 7 symptomatic and 8 asymptomatic SHS-exposed subjects with preserved lung function. Panel G shows representative nasal brushings stained for Keratin (K)-5 (green), p63 (red) and DAPI for nuclei (blue) for an asymptomatic and symptomatic subject. Red arrows indicate K5/p63 dual positive airway stem/progenitor cells. Panel H shows the impact of symptoms on K5/p63 dual positive airway stem/progenitor cells. K5/p63 dual positive airway stem/progenitors significantly correlate with I) total CAT, and trend toward correlation with J) CRP and K) total lung capacity (TLC). P-values indicate results of t-tests for panels A-F and H, where data are shown as mean and 95% CI. Pearson correlation coefficients were estimated for panels I-K.

## DISCUSSION

The World Health Organization estimates that over one third of the world’s adult population is exposed to SHS every year(1), which is almost twice the number of active smokers(44). Although ongoing SHS-exposure has many harmful effects(5, 6), investigations have only touched on the possibility that SHS-exposure might be associated with respiratory disease years after exposure has stopped, or that it may result in newly recognized forms of symptomatic COPD with preserved lung function(45, 46). This prospective, cohort study finds that remote SHS-exposure is associated with worsened respiratory health decades after exposure has ceased. It also found that the presence of respiratory symptoms in SHS-exposed subjects with preserved lung function has both clinical and biologic consequences similar to what has recently been described in symptomatic smokers with preserved lung function(14-22, 40, 47, 48).

Prior studies have shown that active SHS exposure is associated with worsened quality of life(49, 50), increased respiratory symptoms(5, 6) and small decrements in lung function^29,57-60^. Our study strengthens and extends these findings by demonstrating that SHS-exposure was associated with similar effects on quality of life and symptoms almost 3 decades after exposure stopped, but was not associated with the presence of obstructive lung disease. This suggests that, in isolation, SHS may be less likely to cause classical COPD with spirometric evidence of obstruction, except in highly susceptible people, such as those with alpha-1 antitrypsin deficiency or if combined with smoking or other environmental exposures(51-53). Results showing that SHS-exposure synergistically interacted with past smoking to increase total SGRQ support this possibility.

Study results demonstrate that SHS-exposure is associated with symptomatic, non-obstructive respiratory disease in never-smokers, and that symptomatic subjects share a common phenotype found in symptomatic smokers with preserved lung function(14-21). For instance, symptoms were present in up to 33% of SHS-exposed subjects with preserved lung function, which is similar to smokers with preserved lung function where 50% were symptomatic(14, 15). Symptoms in SHS-exposed subjects with preserved lung function were also associated with reduced quality of life, decrements of lung function within the normal range, increased use of inhaled bronchodilators and a prior history of chronic bronchitis, replicating characteristics of symptomatic smokers with preserved lung function(14-21). Study results also support work by Arjomandi *et al*. demonstrating that respiratory complaints were present in 50% of SHS-exposed flight attendants with preserved lung function(46). Together, findings suggest that SHS-exposure is more likely to be associated with non-classical, symptomatic COPD with preserved lung function than to classical COPD with spirometric evidence of obstruction.

SHS-associated respiratory disease also shares a common endotype with symptomatic smokers with preserved lung function(22, 40, 47). For example, inflammatory biomarkers, CRP and SAA-1, were higher in plasma of symptomatic SHS-exposed never smokers with preserved lung function, mirroring findings in a similar population of symptomatic smokers(22). Sinonasal airway basal stem/progenitor cells were also decreased in symptomatic SHS-exposed never-smokers with preserved lung function. These results are similar to findings in smokers where decreased stem/progenitor cells identified subjects with COPD, and non-COPD subjects with intermediate lung function that was in the low normal range(40, 47). Airway basal stem/progenitor cells are critical for airway health, because of their ability to replace injured airway epithelial cells and to maintain homeostasis(47, 48, 54). Accordingly, prior SHS-exposure may have long-term effects on biologic mechanisms that drive development of respiratory disease.

The study has several limitations to consider. The study enrolled exposed and unexposed subjects at a 3:1 ratio, rather than the planned 2:1 ratio, which resulted in 95% power with a nonparametric adjustment for long right tails to detect the 5.6 unit mean difference in total SGRQ using the observed SD of 13.9 in the exposed and 12.3 in the unexposed group. The study was performed primarily in females, which limits generalizability. But it also adds to the limited number of studies that have focused on biological sex differences, and strengthens the notion that SHS-exposure has significant effects in females(49). Symptomatic, SHS-exposed subjects with preserved lung function reported more parental smoking during pregnancy and more home SHS exposure than asymptomatic subjects. This reinforces the concept that cumulative exposure may be important in the development and persistence of respiratory diseases like COPD with a long latency period(12, 32, 55), especially if the exposure occurs during periods of lung development and growth(52, 53, 56, 57). Finally, the control group included unexposed flight attendants and non-flight attendants, making it possible that other exposures unique to flight attendants could have contributed to increased SGRQ in the SHS-exposed group. While we cannot exclude this possibility completely, univariable and multiple regression analyses indicated that exposures, such as VGDF, did not contribute to the increase in SGRQ in the SHS-exposed group.

From the 1930s through the 1990s, predominantly female flight attendants, hired in their early 20s(58), were involuntarily exposed to high concentrations of SHS that were ∼14 times greater than the average person and > 6 times greater than the average worker (Fig. 1B)(30). Exposure of flight attendants to SHS was comparable to exposures that continue to occur today in casinos(59, 60), bars(61-63), nightclubs(61, 62), automobiles(64-66), and in the home(67), making the findings from this study relevant to people living in regions that lack public smoking bans, as well as those who continue to be exposed in family homes and vehicles(68).

Results of this study have important prevention, diagnostic and therapeutic implications for billions of people worldwide who have been, and continue to be, exposed to SHS. They also increase the likelihood that SHS contributes to cases of COPD that are not directly caused by smoking, particularly novel forms of COPD with preserved lung function. Hopefully, a better understanding of the long-term impact of SHS exposure will encourage medical providers to ask important questions about the extent of their patients’ exposure to SHS(69), scientists to explore mechanisms of disease and to test therapies that are sorely needed, and legislative bodies to continue to enact public measures to protect all non-smokers from involuntary exposure to SHS.

## Data Availability

De-identified data from this study will be made available to other investigators upon request.

## ACKNOWLEDGEMENTS

We gratefully acknowledge the enthusiastic support from many flight attendants and flight attendant controls, without which this study would not have been possible. We are also indebted to the Denver Chapter of the Clipped Wings United Airlines Stewardess Alumnae, Inc., the Denver Interline Association of Flight Attendants, Ken Kyle of the Association of Flight Attendants-CWA, Denver Council 09 and Judith Anderson of the Association of Flight Attendants-CWA, AFL-CIO. We also thank Jessica Parker of The Union of Southwest Flight Attendants, Denver Domicile Executive Board, TWU Local 556. Finally a heartfelt thank you goes to the Flight Attendant Medical Research Institute for funding this study, and to the courageous flight attendants who fought the tobacco industry in the courts and in the United States Congress to ban smoking on airlines.

## Author Contributions

Vandivier, Min and Diaz del Valle had full access to all of the data in the study and take responsibility for the integrity of the data and the accuracy of the data analysis.

*Study Design and Concept:* Vandivier

*Acquisition of data:* Diaz del Valle, Zakrajsek and Bell

*Analysis and interpretation of data:* Min, Diaz del Valle, Koff, Zakrajsek, Bell, Kincaid, Frank, Ramakrishnan, Ghosh and Vandivier

*Drafting of the manuscript:* Diaz del Valle, Min, Koff, Ghosh and Vandivier

*Critical revision of the manuscript for important intellectual content:* Diaz del Valle, Zakrajsek, Min, Koff, Kincaid, Frank, Ramakrishnan, Ghosh and Vandivier

*Statistical analysis:* Min and Vandivier

*Administrative, technical, or material support:* Diaz del Valle, Zakrajsek and Vandivier

*Study supervision:* Diaz del Valle and Vandivier

## Online Data Supplement

This article has an online data supplement, which is accessible from this issue’s table of content online at www.atsjournal.org

## REFERENCES

1. Global Estimate of the Burden of Disease From Second-Hand Smoke. 2011 2011. Available from: https://www.who.int/tobacco/publications/second_hand/global_estimate_burden_disease/en/.

2. Quantifying Envorinmental Health Impacts. 2021 [cited 2021 2/12/2021]. Available from: https://www.who.int/quantifying_ehimpacts/publications/shsarticle2010/en/.

3. Homa DM, Neff LJ, King BA, Caraballo RS, Bunnell RE, Babb SD, Garrett BE, Sosnoff CS, Wang L, Centers for Disease C, Prevention. Vital signs: disparities in nonsmokers’ exposure to secondhand smoke--United States, 1999-2012. MMWR Morb Mortal Wkly Rep 2015; 64: 103–108.

4. Max W, Sung HY, Shi Y. Deaths from secondhand smoke exposure in the United States: economic implications. Am J Public Health 2012; 102: 2173–2180.

5. The health consequences of involuntary exposure to tobacco smoke: a report of the Surgeon General. In: U.S. Department of Health and Human Services PHS, editor. Rockville, MD Washington DC; 2006.

6. The Health Consequences of Smoking - 50 Years of Progress. A Report of the Surgeon General. In: U.S. Department of Health and Human Services PHS, editor; 2014. p. 1–943.

7. Global Strategy for the Diagnosis, Managment, and Prevention of Chronic Obstructive Pulmonary Disease. 2021 [cited 2021. Available from: www.goldcopd.org.

8. McDonough JE, Yuan R, Suzuki M, Seyednejad N, Elliott WM, Sanchez PG, Wright AC, Gefter WB, Litzky L, Coxson HO, Pare PD, Sin DD, Pierce RA, Woods JC, McWilliams AM, Mayo JR, Lam SC, Cooper JD, Hogg JC. Small-airway obstruction and emphysema in chronic obstructive pulmonary disease. N Engl J Med 2011; 365: 1567–1575.

9. Hogg JC. A pathologist’s view of airway obstruction in chronic obstructive pulmonary disease. Am J Respir Crit Care Med 2012; 186: v–vii.

10. Hogg JC, McDonough JE, Suzuki M. Small airway obstruction in COPD: new insights based on micro-CT imaging and MRI imaging. Chest 2013; 143: 1436–1443.

11. Lange P, Celli B, Agusti A, Boje Jensen G, Divo M, Faner R, Guerra S, Marott JL, Martinez FD, Martinez-Camblor P, Meek P, Owen CA, Petersen H, Pinto-Plata V, Schnohr P, Sood A, Soriano JB, Tesfaigzi Y, Vestbo J. Lung-Function Trajectories Leading to Chronic Obstructive Pulmonary Disease. N Engl J Med 2015; 373: 111–122.

12. Rennard SI, Drummond MB. Early chronic obstructive pulmonary disease: definition, assessment, and prevention. Lancet 2015; 385: 1778–1788.

13. Agusti A, Celli B. Natural history of COPD: gaps and opportunities. ERJ Open Res 2017; 3.

14. Woodruff PG, Barr RG, Bleecker E, Christenson SA, Couper D, Curtis JL, Gouskova NA, Hansel NN, Hoffman EA, Kanner RE, Kleerup E, Lazarus SC, Martinez FJ, Paine R, 3rd, Rennard S, Tashkin DP, Han MK, Group SR. Clinical Significance of Symptoms in Smokers with Preserved Pulmonary Function. N Engl J Med 2016; 374: 1811–1821.

15. Regan EA, Lynch DA, Curran-Everett D, Curtis JL, Austin JH, Grenier PA, Kauczor HU, Bailey WC, DeMeo DL, Casaburi RH, Friedman P, Van Beek EJ, Hokanson JE, Bowler RP, Beaty TH, Washko GR, Han MK, Kim V, Kim SS, Yagihashi K, Washington L, McEvoy CE, Tanner C, Mannino DM, Make BJ, Silverman EK, Crapo JD, Genetic Epidemiology of CI. Clinical and Radiologic Disease in Smokers With Normal Spirometry. JAMA Intern Med 2015; 175: 1539–1549.

16. Balte PP, Chaves PHM, Couper DJ, Enright P, Jacobs DR, Jr., Kalhan R, Kronmal RA, Loehr LR, London SJ, Newman AB, O’Connor GT, Schwartz JE, Smith BM, Smith LJ, White WB, Yende S, Oelsner EC. Association of Nonobstructive Chronic Bronchitis With Respiratory Health Outcomes in Adults. JAMA Intern Med 2020.

17. Oelsner EC, Balte PP, Bhatt SP, Cassano PA, Couper D, Folsom AR, Freedman ND, Jacobs DR, Jr., Kalhan R, Mathew AR, Kronmal RA, Loehr LR, London SJ, Newman AB, O’Connor GT, Schwartz JE, Smith LJ, White WB, Yende S. Lung function decline in former smokers and low-intensity current smokers: a secondary data analysis of the NHLBI Pooled Cohorts Study. Lancet Respir Med 2020; 8: 34–44.

18. Oelsner EC, Hoffman EA, Folsom AR, Carr JJ, Enright PL, Kawut SM, Kronmal R, Lederer D, Lima JA, Lovasi GS, Shea S, Barr RG. Association between emphysema-like lung on cardiac computed tomography and mortality in persons without airflow obstruction: a cohort study. Ann Intern Med 2014; 161: 863–873.

19. Han MK, Agusti A, Celli BR, Criner GJ, Halpin DMG, Roche N, Papi A, Stockley RA, Wedzicha J, Vogelmeier CF. From GOLD 0 to Pre-COPD. Am J Respir Crit Care Med 2021; 203: 414–423.

20. Martinez FJ, Han MK, Allinson JP, Barr RG, Boucher RC, Calverley PMA, Celli BR, Christenson SA, Crystal RG, Fageras M, Freeman CM, Groenke L, Hoffman EA, Kesimer M, Kostikas K, Paine R, 3rd, Rafii S, Rennard SI, Segal LN, Shaykhiev R, Stevenson C, Tal-Singer R, Vestbo J, Woodruff PG, Curtis JL, Wedzicha JA. At the Root: Defining and Halting Progression of Early Chronic Obstructive Pulmonary Disease. Am J Respir Crit Care Med 2018; 197: 1540–1551.

21. Oh AL, Mularski RA, Barjaktarevic I, Barr RG, Bowler RP, Comellas AP, Cooper CB, Criner GJ, Han MK, Hansel NN, Hoffman EA, Kanner RE, Krishnan JA, Paine R, 3rd, Parekh TM, Peters SP, Christenson SA, Woodruff PG, Investigators S. Defining Resilience to Smoking Related Lung Disease: A Modified Delphi Approach from SPIROMICS. Ann Am Thorac Soc 2021.

22. Garudadri S, Woodruff PG, Han MK, Curtis JL, Barr RG, Bleecker ER, Bowler RP, Comellas A, Cooper CB, Criner G, Dransfield MT, Hansel NN, Paine R, 3rd, Krishnan JA, Peters SP, Hastie AT, Martinez FJ, O’Neal WK, Couper DJ, Alexis NE, Christenson SA. Systemic Markers of Inflammation in Smokers With Symptoms Despite Preserved Spirometry in SPIROMICS. Chest 2019; 155: 908–917.

23. Salvi SS, Barnes PJ. Chronic obstructive pulmonary disease in non-smokers. Lancet 2009; 374: 733–743.

24. Centers for Disease C, Prevention. Chronic obstructive pulmonary disease among adults--United States, 2011. MMWR Morb Mortal Wkly Rep 2012; 61: 938–943.

25. Syamlal G, Doney B, Mazurek JM. Chronic Obstructive Pulmonary Disease Prevalence Among Adults Who Have Never Smoked, by Industry and Occupation - United States, 2013-2017. MMWR Morb Mortal Wkly Rep 2019; 68: 303–307.

26. Wheaton AG, Liu Y, Croft JB, VanFrank B, Croxton TL, Punturieri A, Postow L, Greenlund KJ. Chronic Obstructive Pulmonary Disease and Smoking Status - United States, 2017. MMWR Morb Mortal Wkly Rep 2019; 68: 533–538.

27. Blanc PD, Iribarren C, Trupin L, Earnest G, Katz PP, Balmes J, Sidney S, Eisner MD. Occupational exposures and the risk of COPD: dusty trades revisited. Thorax 2009; 64: 6–12.

28. Blanc PD, Toren K. Occupation in chronic obstructive pulmonary disease and chronic bronchitis: an update. Int J Tuberc Lung Dis 2007; 11: 251–257.

29. Toren K, Jarvholm B. Effect of occupational exposure to vapors, gases, dusts, and fumes on COPD mortality risk among Swedish construction workers: a longitudinal cohort study. Chest 2014; 145: 992–997.

30. Repace J. Flying the smoky skies: secondhand smoke exposure of flight attendants. Tob Control 2004; 13 Suppl 1: i8–19.

31. Abbey DE, Burchette RJ, Knutsen SF, McDonnell WF, Lebowitz MD, Enright PL. Long-term particulate and other air pollutants and lung function in nonsmokers. Am J Respir Crit Care Med 1998; 158: 289–298.

32. Eisner MD, Balmes J, Katz PP, Trupin L, Yelin EH, Blanc PD. Lifetime environmental tobacco smoke exposure and the risk of chronic obstructive pulmonary disease. Environ Health 2005; 4: 7.

33. Arjomandi M, Haight T, Redberg R, Gold WM. Pulmonary function abnormalities in never-smoking flight attendants exposed to secondhand tobacco smoke in the aircraft cabin. J Occup Environ Med 2009; 51: 639–646.

34. Jones PW, Quirk FH, Baveystock CM, Littlejohns P. A self-complete measure of health status for chronic airflow limitation. The St. George’s Respiratory Questionnaire. Am Rev Respir Dis 1992; 145: 1321–1327.

35. Jones PW, Harding G, Berry P, Wiklund I, Chen WH, Kline Leidy N. Development and first validation of the COPD Assessment Test. Eur Respir J 2009; 34: 648–654.

36. Hays RD, Sherbourne CD, Mazel RM. The RAND 36-Item Health Survey 1.0. Health Econ 1993; 2: 217–227.

37. Quanjer PH, Stanojevic S, Cole TJ, Baur X, Hall GL, Culver BH, Enright PL, Hankinson JL, Ip MS, Zheng J, Stocks J, Initiative ERSGLF. Multi-ethnic reference values for spirometry for the 3-95-yr age range: the global lung function 2012 equations. Eur Respir J 2012; 40: 1324–1343.

38. Crapo RO, Morris AH. Standardized single breath normal values for carbon monoxide diffusing capacity. Am Rev Respir Dis 1981; 123: 185–189.

39. Massey CJ, Diaz Del Valle F, Abuzeid WM, Levy JM, Mueller S, Levine CG, Smith SS, Bleier BS, Ramakrishnan VR. Sample collection for laboratory-based study of the nasal airway and sinuses: a research compendium. Int Forum Allergy Rhinol 2020; 10: 303–313.

40. Ghosh M, Miller YE, Nakachi I, Kwon JB, Baron AE, Brantley AE, Merrick DT, Franklin WA, Keith RL, Vandivier RW. Exhaustion of Airway Basal Progenitor Cells in Early and Established Chronic Obstructive Pulmonary Disease. Am J Respir Crit Care Med 2018; 197: 885–896.

41. Jones PW. St. George’s Respiratory Questionnaire: MCID. COPD 2005; 2: 75–79.

42. Kon SS, Canavan JL, Jones SE, Nolan CM, Clark AL, Dickson MJ, Haselden BM, Polkey MI, Man WD. Minimum clinically important difference for the COPD Assessment Test: a prospective analysis. Lancet Respir Med 2014; 2: 195–203.

43. Martinez CH, Murray S, Barr RG, Bleecker E, Bowler RP, Christenson SA, Comellas AP, Cooper CB, Couper D, Criner GJ, Curtis JL, Dransfield MT, Hansel NN, Hoffman EA, Kanner RE, Kleerup E, Krishnan JA, Lazarus SC, Leidy NK, O’Neal W, Martinez FJ, Paine R, 3rd, Rennard SI, Tashkin DP, Woodruff PG, Han MK, Subpopulations, Intermediate Outcome Measures in CSI. Respiratory Symptoms Items from the COPD Assessment Test Identify Ever-Smokers with Preserved Lung Function at Higher Risk for Poor Respiratory Outcomes. An Analysis of the Subpopulations and Intermediate Outcome Measures in COPD Study Cohort. Ann Am Thorac Soc 2017; 14: 636–642.

44. Yousuf H, Hofstra M, Tijssen J, Leenen B, Lindemans JW, van Rossum A, Narula J, Hofstra L. Estimated Worldwide Mortality Attributed to Secondhand Tobacco Smoke Exposure, 1990-2016. JAMA Netw Open 2020; 3: e201177.

45. Flexeder C, Zock JP, Jarvis D, Verlato G, Olivieri M, Benke G, Abramson MJ, Sigsgaard T, Svanes C, Toren K, Nowak D, Jogi R, Martinez-Moratalla J, Demoly P, Janson C, Gislason T, Bono R, Holm M, Franklin KA, Garcia-Aymerich J, Siroux V, Leynaert B, Dorado Arenas S, Corsico AG, Pereira-Vega A, Probst-Hensch N, Urrutia Landa I, Schulz H, Heinrich J. Second-hand smoke exposure in adulthood and lower respiratory health during 20 year follow up in the European Community Respiratory Health Survey. Respir Res 2019; 20: 33.

46. Arjomandi M, Zeng S, Geerts J, Stiner RK, Bos B, van Koeverden I, Keene J, Elicker B, Blanc PD, Gold WM. Lung volumes identify an at-risk group in persons with prolonged secondhand tobacco smoke exposure but without overt airflow obstruction. BMJ Open Respir Res 2018; 5: e000284.

47. Shaykhiev R. Airway Epithelial Progenitors and the Natural History of Chronic Obstructive Pulmonary Disease. Am J Respir Crit Care Med 2018; 197: 847–849.

48. Crystal RG. Airway basal cells. The “smoking gun” of chronic obstructive pulmonary disease. Am J Respir Crit Care Med 2014; 190: 1355–1362.

49. Bridevaux PO, Cornuz J, Gaspoz JM, Burnand B, Ackermann-Liebrich U, Schindler C, Leuenberger P, Rochat T, Gerbase MW, Team S. Secondhand smoke and health-related quality of life in never smokers: results from the SAPALDIA cohort study 2. Arch Intern Med 2007; 167: 2516–2523.

50. Kim YW, Lee CH, Park YS, Kim YI, Ahn CM, Kim JO, Park JH, Lee SH, Kim JY, Chun EM, Jung TH, Yoo KH. Effect of Exposure to Second-Hand Smoke on the Quality of Life: A Nationwide Population-Based Study from South Korea. PLoS One 2015; 10: e0138731.

51. Senn O, Russi EW, Imboden M, Probst-Hensch NM. alpha1-Antitrypsin deficiency and lung disease: risk modification by occupational and environmental inhalants. Eur Respir J 2005; 26: 909–917.

52. Allinson JP, Hardy R, Donaldson GC, Shaheen SO, Kuh D, Wedzicha JA. Combined Impact of Smoking and Early-Life Exposures on Adult Lung Function Trajectories. Am J Respir Crit Care Med 2017; 196: 1021–1030.

53. Bui DS, Perret JL, Walters EH, Abramson MJ, Burgess JA, Bui MQ, Bowatte G, Lowe AJ, Russell MA, Alif SM, Thompson BR, Hamilton GS, Giles GG, Thomas PS, Morrison S, Johns DP, Knibbs LD, Zock JP, Marcon A, Garcia-Aymerich J, Erbas B, Jarvis D, Svanes C, Lodge CJ, Dharmage SC. Lifetime Risk Factors for Pre- and Post-Bronchodilator Lung Function Decline. A Population-based Study. Ann Am Thorac Soc 2020; 17: 302–312.

54. Rock JR, Onaitis MW, Rawlins EL, Lu Y, Clark CP, Xue Y, Randell SH, Hogan BL. Basal cells as stem cells of the mouse trachea and human airway epithelium. Proc Natl Acad Sci U S A 2009; 106: 12771–12775.

55. Jaakkola MS, Samet JM. Occupational exposure to environmental tobacco smoke and health risk assessment. Environ Health Perspect 1999; 107 Suppl 6: 829–835.

56. Martinez FD. Early-Life Origins of Chronic Obstructive Pulmonary Disease. N Engl J Med 2016; 375: 871–878.

57. Spindel ER, McEvoy CT. The Role of Nicotine in the Effects of Maternal Smoking during Pregnancy on Lung Development and Childhood Respiratory Disease. Implications for Dangers of E-Cigarettes. Am J Respir Crit Care Med 2016; 193: 486–494.

58. Ebbert JO, Croghan IT, Schroeder DR, Murawski J, Hurt RD. Association between respiratory tract diseases and secondhand smoke exposure among never smoking flight attendants: a cross-sectional survey. Environ Health 2007; 6: 28.

59. Babb S, McNeil C, Kruger J, Tynan MA. Secondhand smoke and smoking restrictions in casinos: a review of the evidence. Tob Control 2015; 24: 11–17.

60. Repace JL, Jiang RT, Acevedo-Bolton V, Cheng KC, Klepeis NE, Ott WR, Hildemann LM. Fine particle air pollution and secondhand smoke exposures and risks inside 66 US casinos. Environ Res 2011; 111: 473–484.

61. Jones MR, Wipfli H, Shahrir S, Avila-Tang E, Samet JM, Breysse PN, Navas-Acien A, Investigators FBS. Secondhand tobacco smoke: an occupational hazard for smoking and non-smoking bar and nightclub employees. Tob Control 2013; 22: 308–314.

62. Nebot M, Lopez MJ, Gorini G, Neuberger M, Axelsson S, Pilali M, Fonseca C, Abdennbi K, Hackshaw A, Moshammer H, Laurent AM, Salles J, Georgouli M, Fondelli MC, Serrahima E, Centrich F, Hammond SK. Environmental tobacco smoke exposure in public places of European cities. Tob Control 2005; 14: 60–63.

63. Navas-Acien A, Peruga A, Breysse P, Zavaleta A, Blanco-Marquizo A, Pitarque R, Acuna M, Jimenez-Reyes K, Colombo VL, Gamarra G, Stillman FA, Samet J. Secondhand tobacco smoke in public places in Latin America, 2002-2003. JAMA 2004; 291: 2741–2745.

64. Sendzik T, Fong GT, Travers MJ, Hyland A. An experimental investigation of tobacco smoke pollution in cars. Nicotine Tob Res 2009; 11: 627–634.

65. Semple S, Apsley A, Galea KS, MacCalman L, Friel B, Snelgrove V. Secondhand smoke in cars: assessing children’s potential exposure during typical journey conditions. Tob Control 2012; 21: 578–583.

66. Rees VW, Connolly GN. Measuring air quality to protect children from secondhand smoke in cars. Am J Prev Med 2006; 31: 363–368.

67. Zhang T, Chillrud SN, Yang Q, Pitiranggon M, Ross J, Perera F, Ji J, Spira A, Breysse PN, Rodes CE, Miller R, Yan B. Characterizing peak exposure of secondhand smoke using a real-time PM2.5 monitor. Indoor Air 2020; 30: 98–107.

68. Agaku IT, Odani S, King BA, Armour BS. Prevalence and correlates of secondhand smoke exposure in the home and in a vehicle among youth in the United States. Prev Med 2019; 126: 105745.

69. Klein JD, Chamberlin ME, Kress EA, Geraci MW, Rosenblatt S, Boykan R, Jenssen B, Rosenblatt SM, Milberger S, Adams WG, Goldstein AO, Rigotti NA, Hovell MF, Holm AL, Vandivier RW, Croxton TL, Young PL, Blissard L, Jewell K, Richardson L, Ostrow J, Resnick EA. Asking the Right Questions About Secondhand Smoke. Nicotine Tob Res 2021; 23: 57–62.

