## Supplement for "Long-term Impact of Prior Secondhand Tobacco Smoke Exposure on Respiratory Health": medRxiv Supplement Final.pdf

#### Contents of Online Supplement

**Supplement Table 1.** Univariable and Multivariable Analysis for Total SGRQ

**Supplement Table 2.** Demographics of the Never Smoker Cohort

**Supplement Figure 1.** Impact of Secondhand Tobacco Smoke (SHS) Exposure on General Quality of Life and Lung Function in the Total Cohort.

**Supplement Figure 2.** Impact of Secondhand Tobacco Smoke (SHS) Exposure on Respiratory Health in the Never Smoker Cohort.

**Supplement Figure 3.** Impact of Secondhand Tobacco Smoke (SHS) Exposure on General Quality of Life and Lung Function in the Never Smoker Cohort.

**Supplement Figure 4**. Synergistic Interaction Between Secondhand Tobacco Smoke (SHS) Exposure and Former Smoking in the Total Cohort.

**Supplement Figure 5.** Impact of Symptoms on Lung Function in the Secondhand Tobacco Smoke (SHS) Exposed, Preserved Lung Function Cohort.

**Supplement Figure 6.** Impact of Symptoms on Sinonasal Airway Inflammatory and Ciliated Cells in the Secondhand Tobacco Smoke (SHS) Exposed, Preserved Lung Function Cohort.

**Supplement Table 1. Univariable and Multivariable Analysis for Total SGRQ**

| Quantile Regression (Median) for Total SGRQ | | | | | | | |
|---|---|---|---|---|---|---|---|
| Univariable Analysis | | | | Multiple Regression Analysis | | | |
| **Variable** | **Beta** | **95% CI** | **P-Value** | **Variable** | **Beta** | **95% CI** | **P-Value** |
| Airline Work SHS-Exposure | 6.7 | 2.7, 10.7 | 0.001 | Airline Work SHS-Exposure | 6.5 | 3.7, 9.2 | <0.001 |
| Age | 0.4 | 0.1, 0.7 | 0.003 | Age | | | NS |
| Female | 1.2 | -4.1, 6.6 | 0.65 | Female | | | NS |
| BMI | 0.8 | 0.3, 1.3 | <0.001 | BMI | 0.8 | 0.5, 1.1 | <0.001 |
| Parental Smoking During Pregnancy | 2.6 | -1.1, 6.4 | 0.17 | Parental Smoking During Pregnancy | 2.7 | 0.3, 5.1 | 0.03 |
| Non-Airline Work SHS-Exposure | 0.1 | -0.0009, 0.3 | 0.07 | Non-Airline SHS-Exposure | | | NS |
| Workplace VGDF-Exposure | 0.04 | -0.1, 0.1 | 0.53 | Workplace VGDF-Exposure | | | NS |
| HTN | 1.1 | -4.0, 6.2 | 0.67 | HTN | | | NS |

*Definition of abbreviations:* SGRQ = St. George’s Respiratory Questionnaire, CI = confidence interval, SHS = secondhand cigarette smoke, BMI = body mass index, VGDF = (self-reported exposure to) vapors, gases, dusts or fumes, HTN = hypertension, NS = non-significant.

**Supplement Table 2. Demographics of the Never Smokers in the Total Cohort***

| Characteristics | SHS Exposed N=131 | SHS Unexposed N=42 | P-value† |
|---|---|---|---|
| Age, y | 66.0 (7.2) | 63.0 (7.6) | 0.02 |
| Gender, n (%) female | 118 (90.1) | 32 (76.2) | 0.02 |
| Race/Ethnicity | | | |
| White, n (%) | 113 (95.8) | 38 (92.7) | 0.43 |
| Black, n (%) | 0 (0) | 0 (0) | |
| Asian, n (%) | 2 (1.7) | 0 (0) | |
| Hispanic, n (%) | 2 (1.7) | 2 (4.9) | |
| Native Hawaiian or Pacific Islander, n (%) | 0 (0) | 0 (0) | |
| Multirace, n (%) | 1 (0.9) | 1 (2.4) | |
| BMI, units | 25.2 (4.1) | 26.4 (4.5) | 0.11 |
| Resting $SpO_2$, % | 95.7 (2.1) | 95.6 (1.9) | 0.74 |
| **Flight Attendant History** | | | |
| Age of Hire, y | 24.0 (4.7) | NA | NA |
| Total Flying Years, y | 29.1 (14.4) | NA | NA |
| Years Elapsed Since SHS-Exposure, y | 27.4 (9.5) | NA | NA |
| **SHS and VGDF Exposure History** | | | |
| Prenatal SHS Exposure | | | 0.31 |
| Only Mother Smoked, n (%) | 6 (4.6) | 2 (4.8) | |
| Only Father Smoked, n (%) | 37 (28.2) | 14 (33.3) | |
| Both Parents Smoked, n (%) | 30 (22.9) | 14 (33.3) | |
| Neither Parent Smoked, n (%) | 58 (44.3) | 12 (28.6) | |
| Parental Smoking During Pregnancy, n (%) | 73 (55.7) | 30 (71.4) | 0.07 |
| Home SHS Exposure, y | 13.2 (11.2) | 13.5 (8.9) | 0.88 |
| Median [IQR] | 17 [0, 20] | 18 [0, 19] | 0.95 |
| Airline Work SHS Exposure, y | 15.3 (9.3) | NA | NA |
| Non-Airline Work SHS Exposure, y | 4.5 (7.8) | 18.5 (16.8) | <0.001 |
| Median [IQR] | 1 [0, 5] | 19.5 [0, 37] | <0.001 |
| Workplace VGDF Exposure, y | 10.4 (15.7) | 3.3 (7.8) | <0.001 |
| Median [IQR] | 0 [0, 26] | 0 [0, 0] | 0.03 |
| **Comorbidities** | | | |
| HTN, n (%) | 36 (27.5) | 11 (26.2) | 0.87 |
| CHF, n (%) | 0 (0) | 1 (2.4) | 0.24 |
| CAD, n (%) | 4 (3.1) | 1 (2.4) | >0.99 |
| DM, n (%) | 2 (1.5) | 4 (9.5) | 0.03 |
| High Cholesterol, n (%) | 39 (29.8) | 9 (21.4) | 0.29 |
| Chronic Bronchitis, n (%) | 6 (4.6) | 2 (4.8) | >0.99 |
| Emphysema, n (%) | 1 (0.8) | 1 (2.4) | 0.43 |
| Asthma, n (%) | 12 (9.2) | 6 (14.3) | 0.34 |
| Sinus Problems, n (%) | 43 (32.8) | 10 (23.8) | 0.27 |
| Ear Infections, n (%) | 13 (9.9) | 2 (4.8) | 0.53 |
| Osteopenia/Osteoporosis, n (%) | 24 (18.3) | 6 (14.3) | 0.55 |
| Sleep Apnea, n (%) | 11 (8.4) | 5 (11.9) | 0.49 |
| Depression, n (%) | 19 (14.5) | 9 (21.4) | 0.29 |
| Anxiety, n (%) | 20 (15.3) | 9 (21.4) | 0.35 |
| Thyroid Disease, n (%) | 25 (19.1) | 10 (23.8) | 0.51 |
| Skin Cancer, n (%) | 15 (11.5) | 3 (7.1) | 0.57 |

| | | | |
|---|---|---|---|
| Total Non-skin Cancer, n (%) | 20 (15.3) | 3 (7.1) | 0.29 |

*Data are presented as mean (SD) unless otherwise stated. Percentages may not total 100% due to rounding. There were no missing data, except 8% for race/ethnicity.

*Definition of abbreviations:* SHS = secondhand cigarette smoke, BMI = body mass index, IQR = interquartile range, $SpO_2\%$ = peripheral blood oxygen saturation, VGDF = (self-reported exposure to) vapors, gases, dusts or fumes, HTN = hypertension, CHF = congestive heart failure and CAD = coronary artery disease, DM = diabetes mellitus

†T-tests (or Mann-Whitney tests) were used for continuous variables and chi-square tests (or Fisher's exact tests) were used for categorical variables.

### Supplement Figure 1

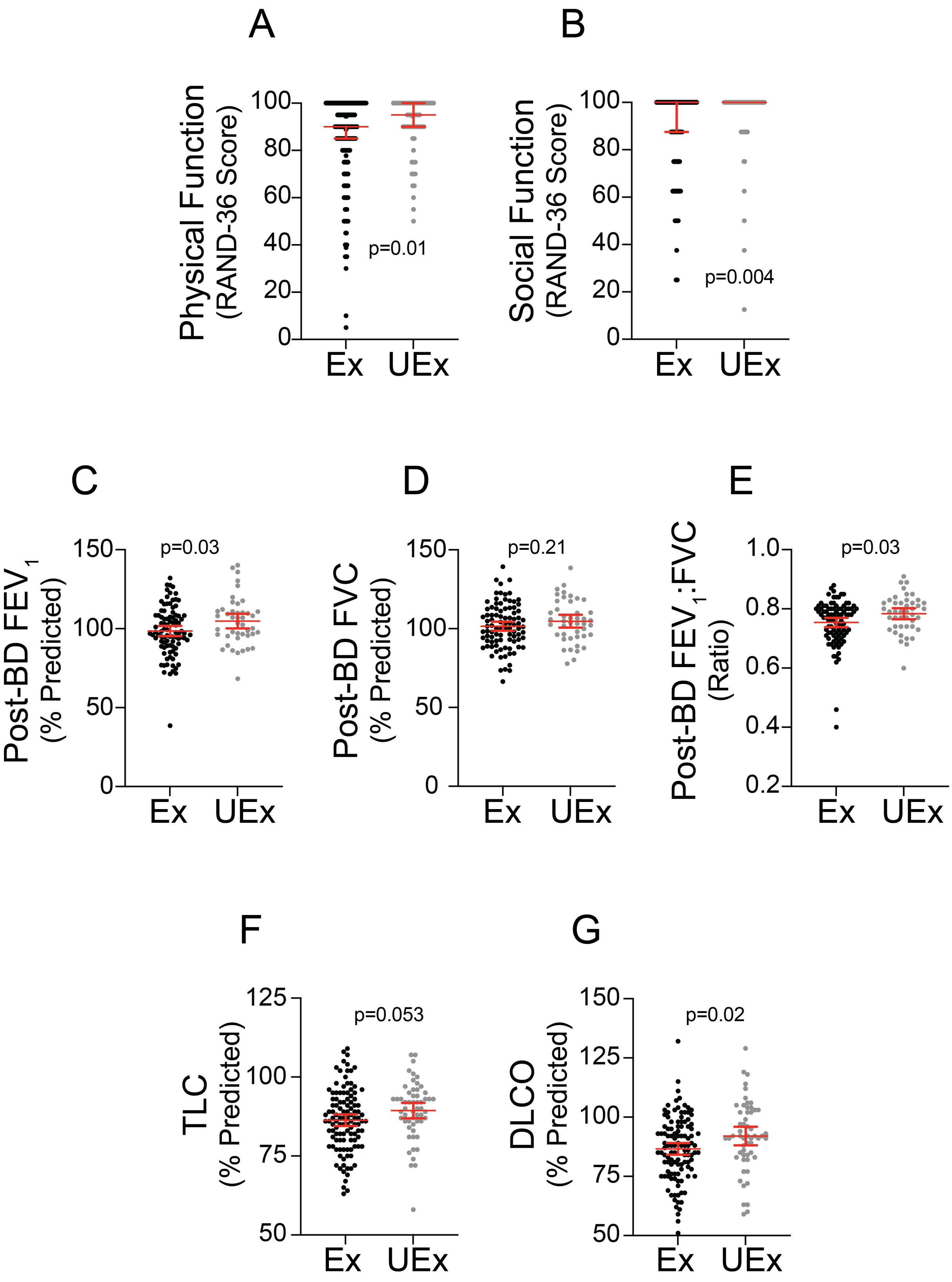

Supplement Figure 2

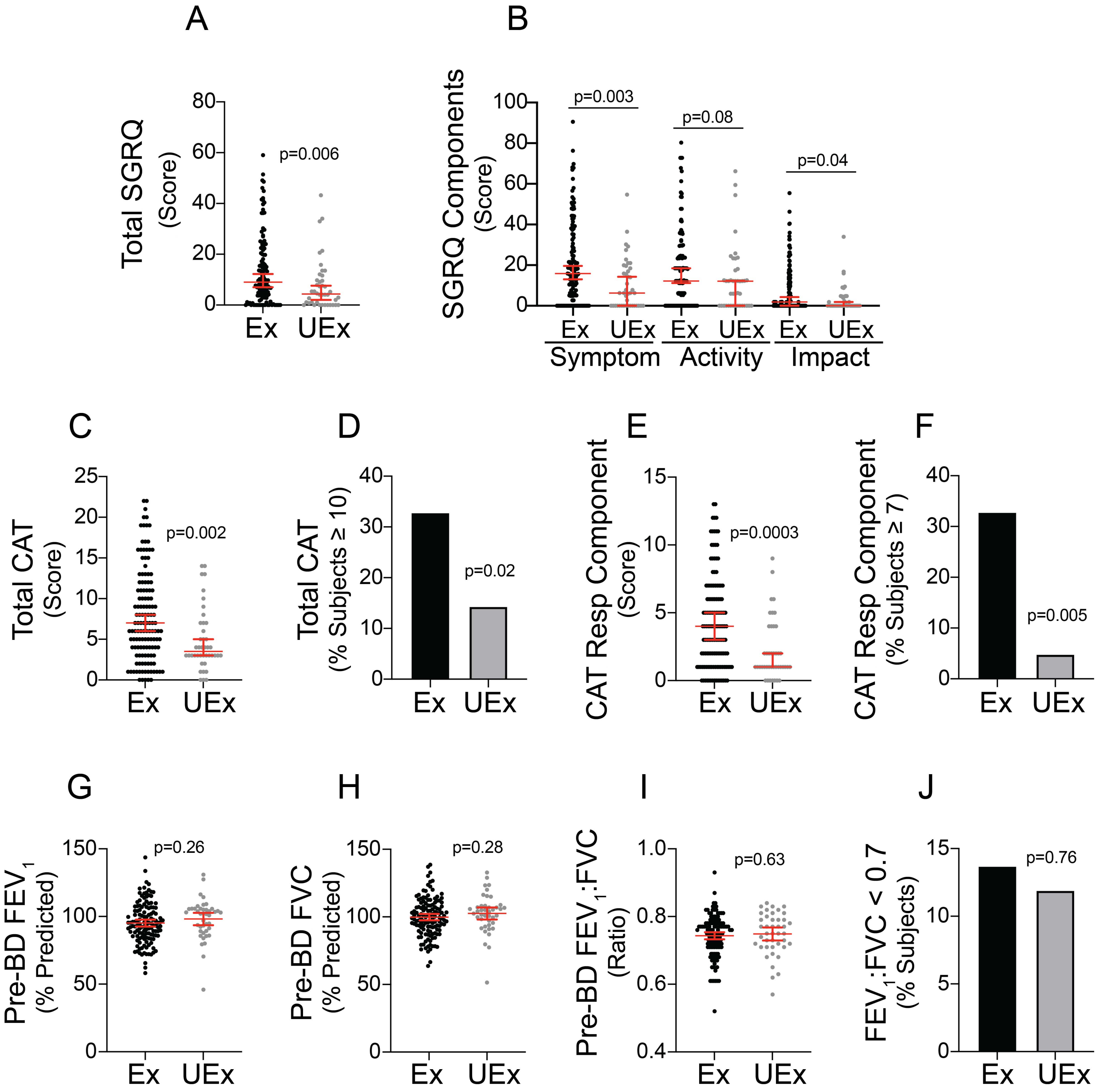

### Supplement Figure 3

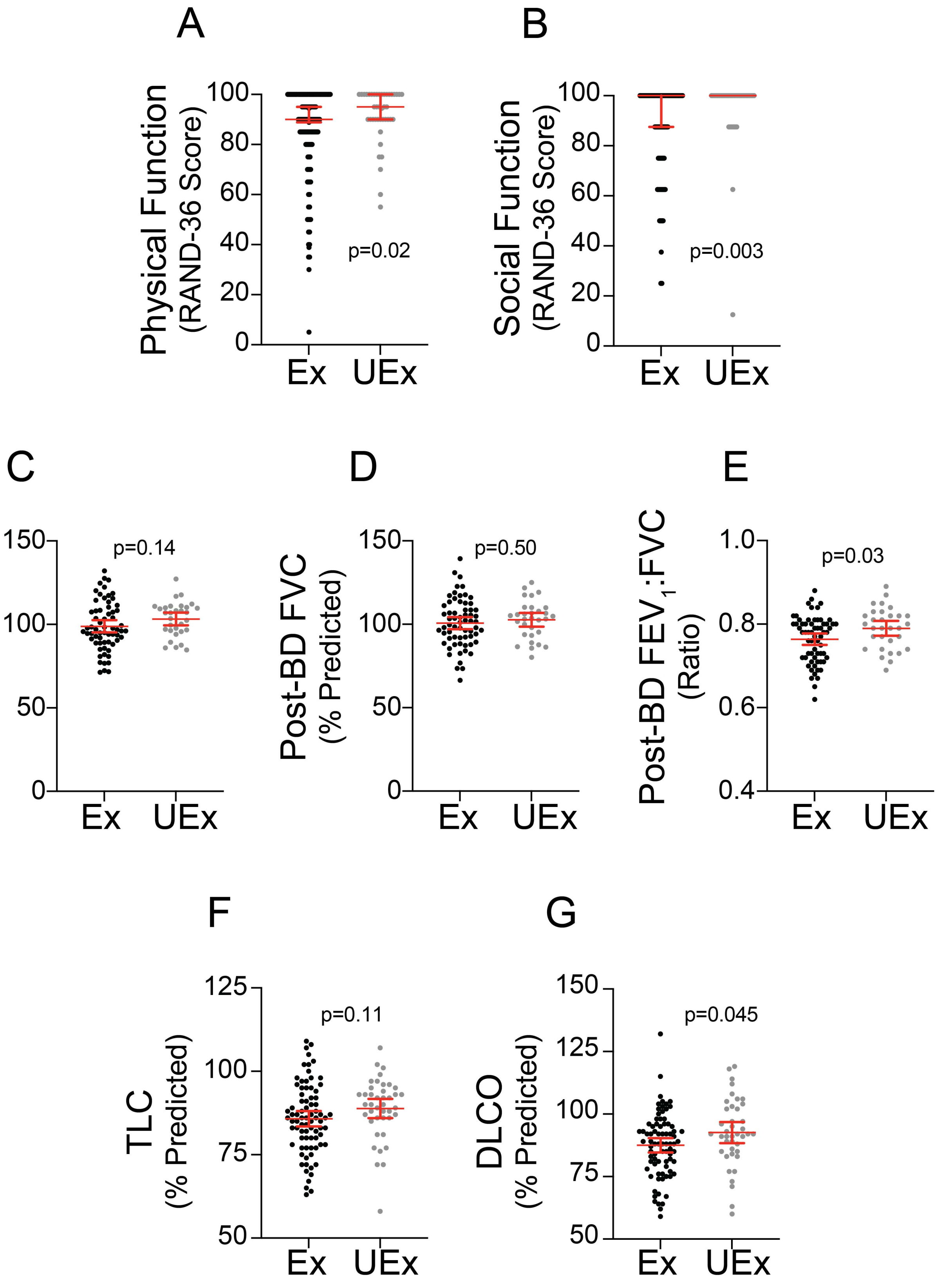

Supplement Figure 4

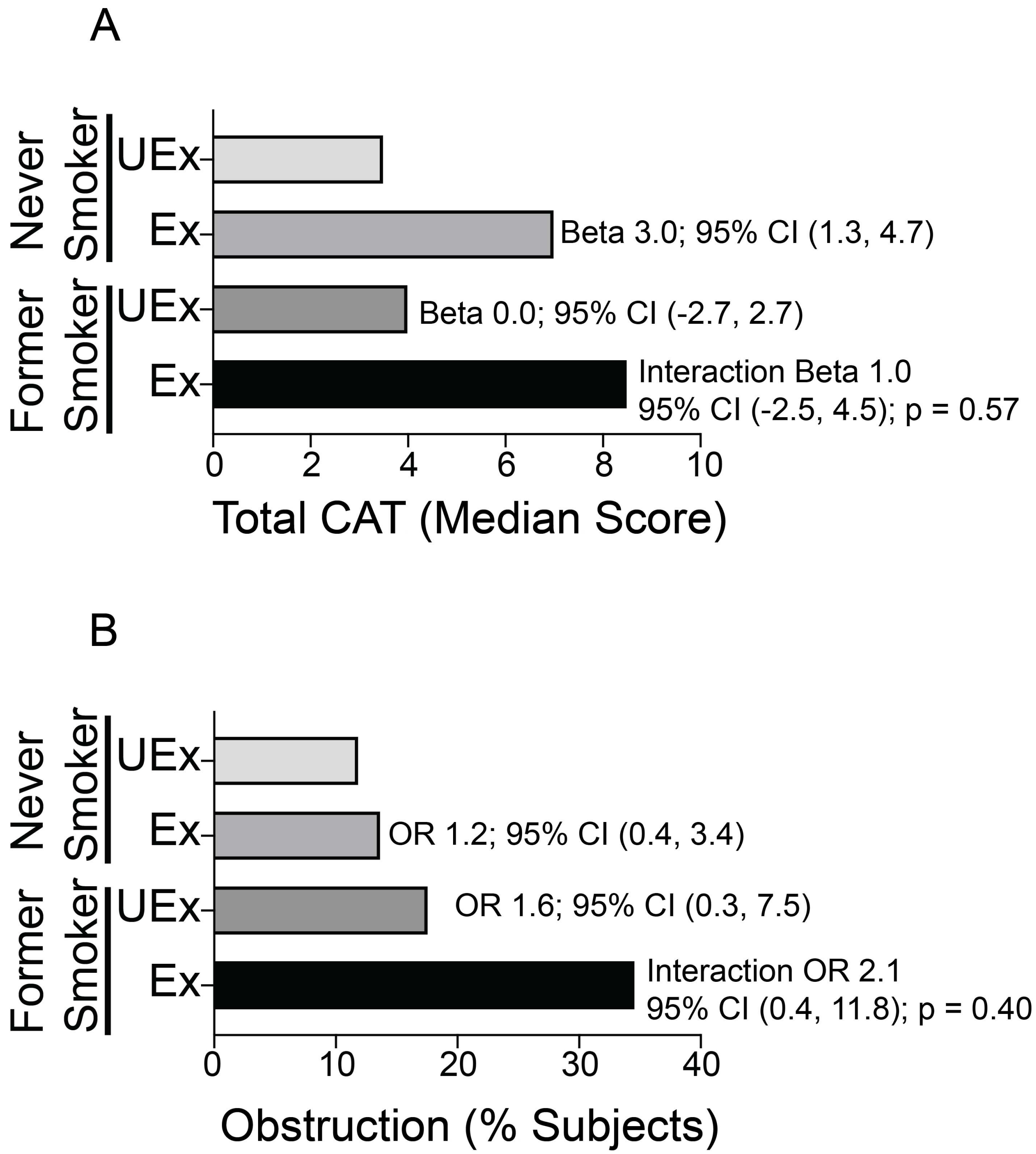

Supplement Figure 5

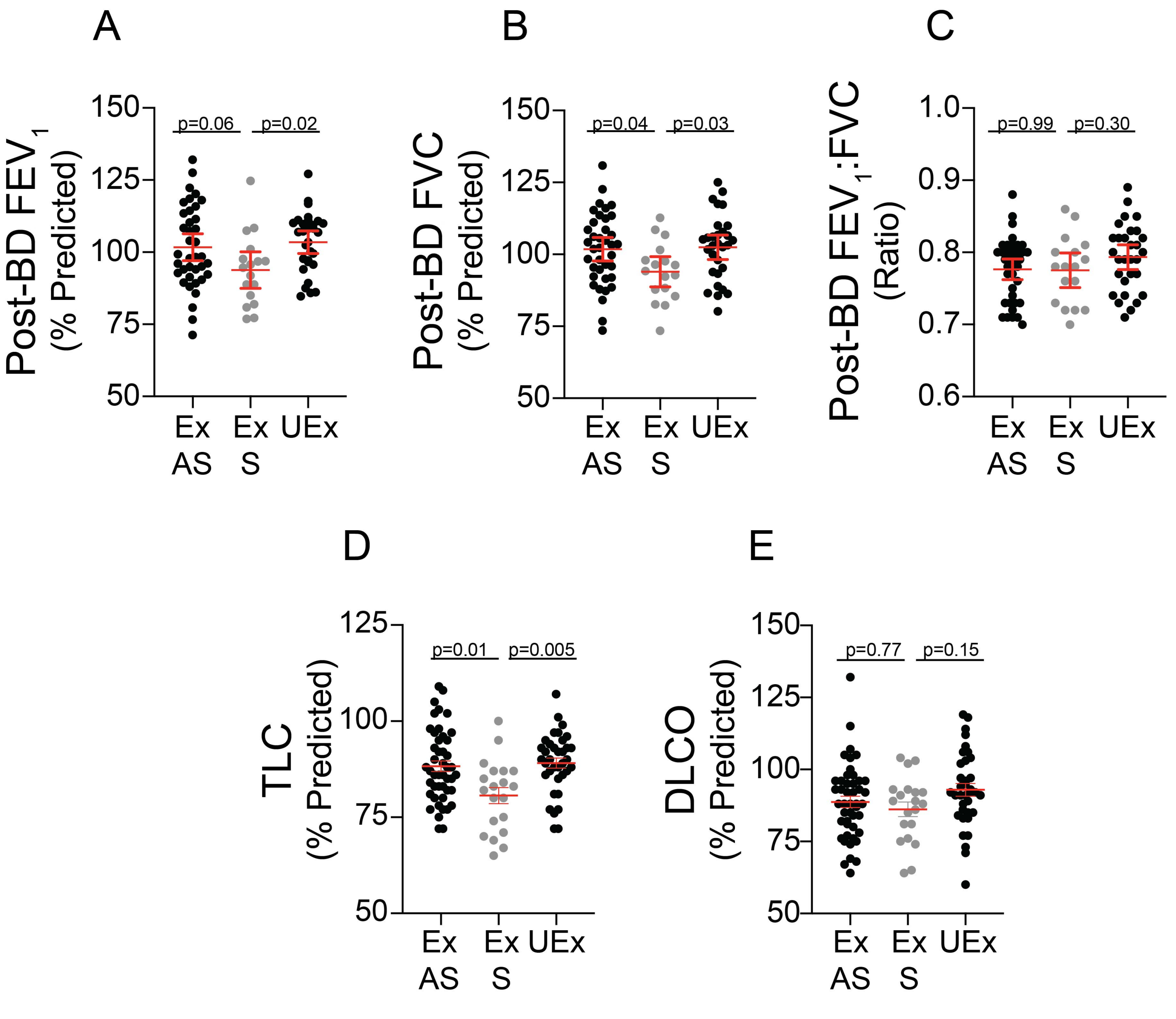

Supplement Figure 6

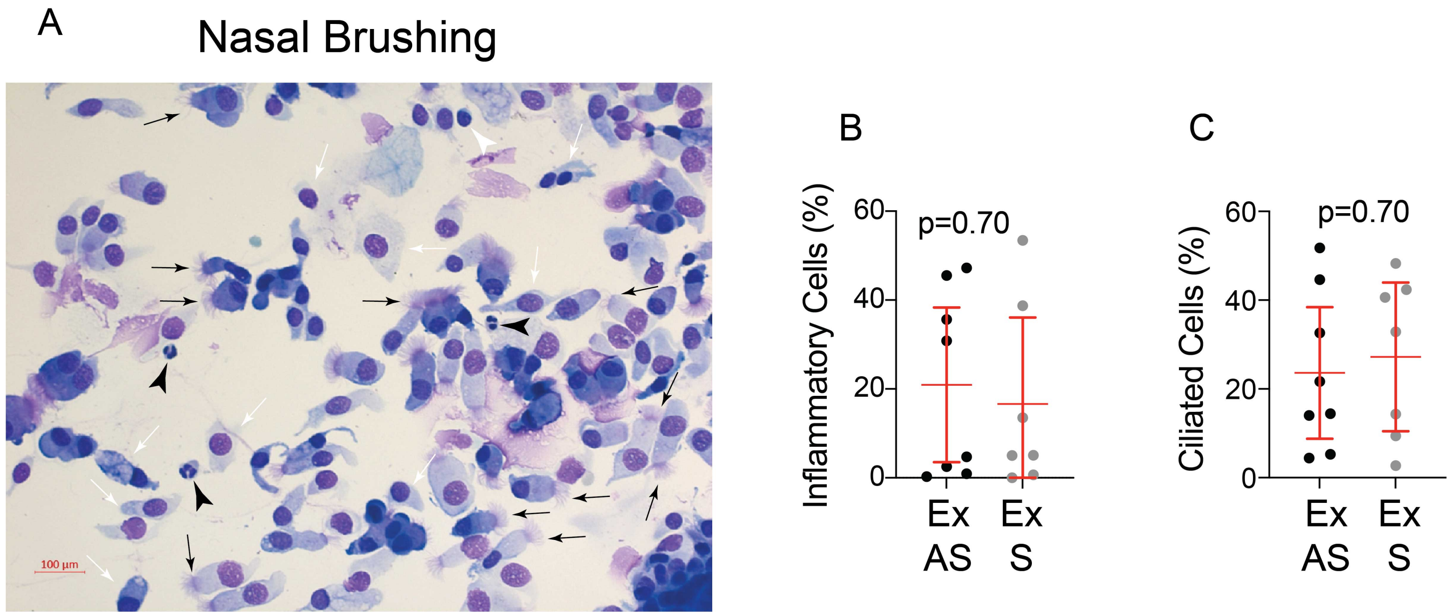

**FIGURE LEGENDS**

**Supplement Figure 1. Impact of Secondhand Tobacco Smoke (SHS) Exposure on General Quality of Life and Lung Function in the Total Cohort.** The study examined differences in SHS-exposed (Ex; N=183) *versus* unexposed (UEx; N=59) subjects for the A) physical function and B) social function components of the RAND Corporation modification of the short form (SF)-36 questionnaire (RAND-36), post-bronchodilator (post-BD) C) $FEV_1$, D) FVC, and E) $FEV_1$:FVC ratio, F) total lung capacity (TLC), and G) lung diffusion of carbon monoxide (DLCO). P-values indicate results of a Mann-Whitney test for A-B, and data are shown as median and 95% CI. P-values indicate results of a t-test for C-G, and data are shown as mean and 95% CI.

**Supplement Figure 2. Impact of Secondhand Tobacco Smoke (SHS) Exposure on Respiratory Health in the Never Smoker Cohort.** The study examined differences in SHS-exposed (Ex; N=131) *versus* unexposed (UEx; N=42) never smokers for A) total St. George's Respiratory Questionnaire (SGRQ), B) symptom, activity and impact components of the SGRQ, C) total COPD Assessment Test (CAT), D) CAT ≥ 10, E) CAT Respiratory (Resp) Component, F) CAT Resp Component ≥ 7, pre-bronchodilator (pre-BD) G) $FEV_1$, H) FVC, and I) $FEV_1$:FVC ratio, and J) obstruction defined by an $FEV_1$:FVC ratio < 0.7 using post-BD measurements where available. P-values indicate results of a Mann-Whitney test for A-C and E, and data are shown as median and 95% CI. P-values indicate results of a t-test for G-I, and data are shown as mean and 95% CI. P-values indicate results of a Fisher's exact test for D, F and J.

**Supplement Figure 3. Impact of Secondhand Tobacco Smoke (SHS) Exposure on General Quality of Life and Lung Function in the Never Smoker Cohort.** The study examined differences in SHS-exposed (Ex; N=131) *versus* unexposed (UEx; N=42) never-smokers for the A) physical function and B) social function components of the RAND Corporation modification of the short form (SF)-36 questionnaire (RAND-36), post-bronchodilator (post-BD) C) $FEV_1$, D) FVC, and E) $FEV_1$:FVC ratio, and F) total lung capacity (TLC), and G) lung diffusion of carbon monoxide (DLCO). P-values indicate results of a Mann-Whitney test for A-B, and data are shown as median and 95% CI. P-values indicate results of a t-test for C-G, and data are shown as mean and 95% CI.

**Supplement Figure 4. Synergistic Interaction Between Secondhand Tobacco Smoke (SHS) Exposure and Former Smoking in the Total Cohort.** The study examined whether SHS-exposure (Ex) and former smoking interacted in a synergistic way compared to unexposed (UEx) never smokers for A) total COPD Assessment Test (CAT) and B) obstruction defined by an $FEV_1$:FVC ratio < 0.7 using post-bronchodilator measurements where available. Beta or Odds Ratio (OR) and 95% CI indicate the main and interaction effects of Ex and former smoking using quantile regression for A, and logistic regression for B.

**Supplement Figure 5. Impact of Symptoms on Lung Function in the Secondhand Tobacco Smoke (SHS) Exposed, Preserved Lung Function Cohort.** The study examined the impact of symptoms in secondhand tobacco smoke (SHS) exposed (Ex), never-smokers with preserved lung function. Symptomatic (S; CAT ≥ 10) SHS-subjects with preserved lung function (N=36) were compared to asymptomatic (AS; CAT < 10) SHS-exposed subjects (N=74) and to unexposed (UEx) never-smokers with preserved lung function (N=36) for post-bronchodilator (post-BD) A) $FEV_1$, B) FVC, and C) $FEV_1$:FVC ratio, D) total lung capacity (TLC), and E) lung diffusion of carbon monoxide (DLCO). Differences were assessed using a one-way ANOVA for A-E, which resulted in a $p < 0.05$ for A, B and D. P-values indicate results of a Dunnett's test and data are shown as mean and 95% CI.

**Supplement Figure 6. Impact of Symptoms on Sinonasal Airway Inflammatory and Ciliated Cells in the Secondhand Tobacco Smoke (SHS) Exposed, Preserved Lung Function Cohort.** Panel A shows a representative Diff-Quick-stained cytospin of a nasal brushing demonstrating the presence of ciliated (black arrows) and non-ciliated (white arrows) epithelial cells, neutrophils (black arrowheads) and lymphocytes (white arrowheads). Panels B and C show differences in B) inflammatory cells (neutrophils and lymphocytes) and C) ciliated cells in symptomatic (S; CAT ≥ 10) and asymptomatic (AS; CAT < 10) secondhand tobacco smoke exposed (Ex), never-smokers with preserved lung function. P-values indicate results of t-tests for panels B and C, where data are shown as mean and 95% CI.
